# T cell immunity predicts clinical outcomes on stopping antiretroviral treatment after HIV-specific broadly neutralising antibody therapy

**DOI:** 10.64898/2026.02.02.26345374

**Authors:** Mohammed Altaf, Carla Nel, Timothy Tipoe, Julia Edgar, Panagiota Zacharopoulou, Devinder Srai, Chanice Knight, Ming Lee, Louise-Rae Cherrill, Emanuela Falaschetti, Ane Ogbe, Stephen Fletcher, Hanna Box, Tamara Elliott, Sabine Kinloch, Julie Fox, Amanda Clarke, Sarah Pett, Simon Collins, Maathini Balachandran, Katie Topping, Louise Terry, Kelly Seaton, Georgia Tomaras, Alison Uriel, Chloe Orkin, Kyle Ring, Gary Whitlock, Marta Boffito, Rebecca Sutherland, Ole S. Søgaard, Jesper D. Gunst, Helen Brown, Nicola Robinson, Gabriella Lindegard, Philip Goulder, Graham Taylor, Marina Caskey, Michel Nussenzweig, Sarah Fidler, John Frater, The RIO Study Team

## Abstract

There is no readily accessible, scalable cure for HIV infection. Trials of HIV-specific broadly neutralising antibodies (bNAbs) demonstrate inhibition of viral replication and reduction of the reservoir of latently-infected cells, potentially offering new strategies for HIV eradication. Animal and human studies suggest bNAbs have multiple activities, including a direct antiviral action and a secondary induction of T cell responses, the ‘vaccinal effect’. The RIO trial assessed HIV-specific cell-mediated immunity after dosing with two long-acting bNAbs (10-1074-LS and 3BNC117-LS) in people treated with antiretroviral therapy (ART) since early stage HIV followed by treatment interruption. BNAbs resulted in sustained viral suppression with 75% of participants controlling off ART after 20 weeks. Here we show that HIV-specific T cell immunity was enhanced for at least 36 weeks after bNAbs in aviraemic participants. Gag-specific T cell immune responses predicted virological outcomes. Baseline CD8+ AIM responses predicted longer times to rebound; baseline CD8+ proliferative responses were additionally protective in participants without baseline bNAb resistance mutations. Baseline ELISpot responses were associated with faster rebound. These data highlight the complex interplay between bNAbs and T cells, identify a post-bNAb protective T cell-driven vaccinal effect, and reinforce the role of immune-based interventions as part of HIV cure strategies.

## Main

Despite the dramatic impact of antiretroviral therapy (ART) on HIV-associated mortality and morbidity, there is no cure for HIV infection^1^. Longer-acting antiviral therapies may alleviate some of the burden of daily pill-taking but are not widely available globally. ART presents many challenges around drug resistance, monitoring, stigma and non-AIDS morbidity, underpinning the need for an intervention that can induce sustained ART-free virological remission^2^. Animal studies and small human clinical trials demonstrate that certain HIV-specific broadly neutralising antibodies (bNAbs) can sustain viral control in the absence of ART^3–6^, and that this control may be extended by bNAb-induced protective T cell responses – the ‘vaccinal effect’^7–11^

The bNAbs 3BNC117^12^ and 10-1074^13^ can neutralise a broad diversity of HIV strains and engage Fc-mediated immune effector mechanisms. Human studies have shown control of viraemia following bNAb infusions, with additional evidence for a decrease in the numbers of latently-infected cells comprising the HIV reservoir, the primary target for achieving HIV cure^6^. In addition, three clinical studies have reported enhanced T cell immunity in virally-suppressed participants after bNAb dosing^7–9,11^, consistent with a “vaccinal effect”. In Rhesus macaque models this post-bNAb T cell vaccinal effect has been linked to sustained viral control, and possibly even cure^5,10^. Whether the same potential exists in humans remains unclear.

The RIO study was the first phase II double-blinded randomised controlled trial of dual long-acting (LS) bNAbs (10-1074-LS and 3BNC-117LS) in people with treated primary or early HIV infection (defined as nadir CD4 count >500 cells/µL)^14^. Sixty-eight participants were randomised 1:1 to receive bNAbs (Arm A) or placebo (Arm B) and then undertake an analytical treatment interruption (ATI). BNAb recipients were 91% less likely to rebound compared to placebo 20 weeks after ATI following a single LS-bNAb dose, and remained 78% less likely to rebound by 96 weeks, receiving up to two doses^15^.

Although a direct antiviral effect was expected, here we asked whether participants receiving bNAbs showed a T cell vaccinal effect with associated viral control beyond expected neutralising plasma bNAb levels. We measured T cell responses using multiple approaches and investigated the contribution of T cells pre- and post-bNAb dosing to virological control. Using a combination of T cell assays and clinical outcome data, we show that T cell immunity is associated with viral control after bNAb dosing in people with treated early HIV infection undertaking an antiretroviral treatment interruption.

### Sustained control of HIV viraemia off ART after bNAb dosing

Thirty-four male participants (median age 36, range 31-44 years) were enrolled and randomly allocated into RIO Arm A (**Table 1**; **Figure 1a**). ART was interrupted 2 days after the first infusion of two LS-bNAbs, and plasma viral load was monitored initially weekly and subsequently fortnightly and 4-weekly in those remaining undetectable. Immunological assessments were made at baseline (Week 0) and at Weeks 12, 24 and 36 after bNAb dosing, unless a participant opted into the second dose sub-study. Of 34 participants, 17 (50%) received an optional second dose of dual LS-bNAbs, a median of 27 weeks (range 21 – 50 weeks) after randomisation. In this case the Week 24 sample collection was replaced with a collection at Week 20 prior to the second bNAb dosing. For the analysis, samples from Weeks 20 and 24 are grouped together. Of the 34 participants in Arm A who received dual LS-bNAbs, 75% (n=26) had not met the trial rebound criteria (defined as either two consecutive VL measurements >100,000 copies/mL, or six consecutive weeks >1,000 copies/mL) by Week 20 compared to 11% in the placebo arm. Control off ART was sustained in those receiving bNAbs, with 25% (n=7) not rebounding by Week 96^15^. We hypothesised that this extended control in a subgroup of Arm A participants - in particular those who had not rebounded by 96 weeks - was associated with altered HIV-specific immunity as plasma bNAb concentrations were below the estimated threshold at which viral neutralisation would be expected.

**Figure 1.**
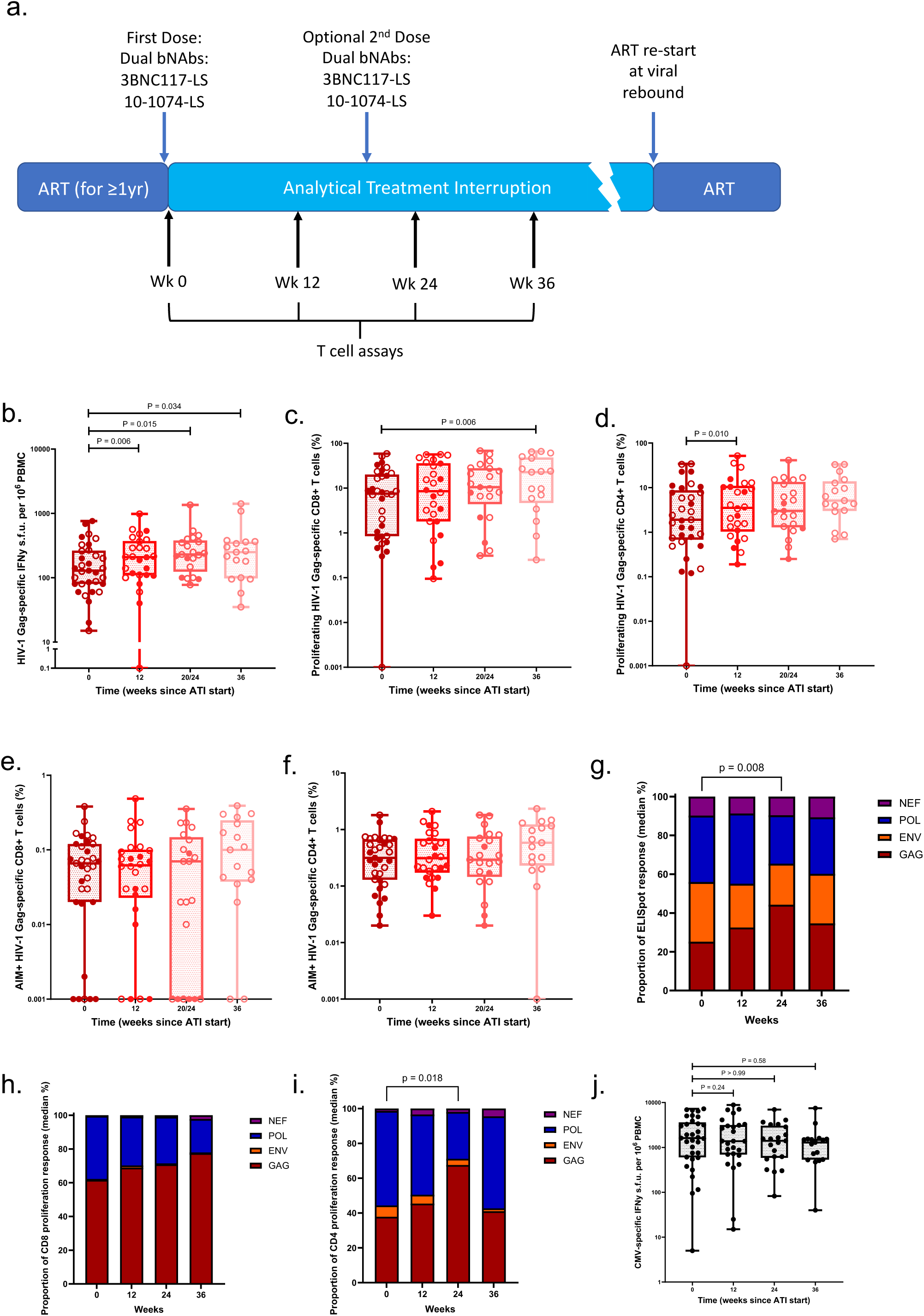
(a) Trial structure and sampling timepoints for RIO Arm A. (b-f) HIV Gag-specific ELISpot, Proliferation (CD8+ and CD4+) and AIM (CD8+ and CD4+) assays 0, 12, 24 and 36 weeks after bNAb dosing and ATI in aviraemic participants. Relative proportions of HIV Gag, Pol, Nef and Env specific responses in (g) ELISpot and Proliferation assays ((h) CD8+ and (i) CD4+); statistical results relate to changes in Gag responses. (j) CMV ELISpot responses after bNAb dosing. IFNy interferon gamma; s.f.u spot forming units; ATI Analytical treatment interruption.

**Table 1.** Demographics. Demographics of participants in Arm A of RIO included in the immunology analysis. IQR Interquartile range; ART antiretroviral therapy; ATI Analytical Treatment Interruption; NRTI Nucleoside Reverse Transcriptase Inhibitor; INSTI Integrase Strand Transfer Inhibitor; bPI boosted Protease Inhibitor.

| **Variable** | **(n=34)** |
| --- | --- |
| **Age** |  |
| Median (IQR) | 36 (31-44) |
| **Sex assigned at birth** |  |
| Male, n (%) | 34 (100%) |
| **Ethnicity, n (%)** |  |
| Asian or Asian British | 3 (8.8%) |
| Black or Black British | 1 (2.9%) |
| White | 28 (82.4%) |
| Other, n (%) | 2 (5.9%) |
| **Duration of ART prior to enrolment, (years)** |  |
| Median (IQR) | 5 (3-6.5) |
| **ART regime at screening, n (%)** |  |
| **NRTI** | 34 (100%) |
| **INSTI** | 27 (79%) |
| **bPI** | 3 (9%) |
| **Baseline CD4+ T cell count, cells/ul** |  |
| Median (IQR) | 742 (657-937) |
| **HIV subtype, n (%)** |  |
| B | 19 (55.9%) |
| Non-B | 6 (17.6%) |
| Unknown | 9 (26.5%) |
| **Stage of HIV at time of diagnosis, n (%)** |  |
| Primary HIV | 32 (94%) |
| Early HIV | 2 (6%) |
| **Human leukocyte antigen class I alleles** |  |
| Susceptible only: B*07:02, B*08:01, B*35, n (%) | 11 (32.4%) |
| Protective only: A*25:01, B*13:02, B*27:05, B*52:01, B*57:01, n (%) | 6 (17.6%) |
| Both susceptible and protective HLA alleles, n (%) | 6 (17.6%) |
| Neither susceptible nor protective HLA alleles, n (%) | 11 (32.4%) |

### HIV Gag-specific T-cell responses increase after LS-bNAbs and an ATI

We explored whether those participants who controlled viraemia off ART after bNAbs showed evidence of new or enhanced T cell immunity. To identify such a ‘vaccinal effect’, T cell responses were quantified at baseline (pre-bNAbs, n=33), and after one dose of bNAbs at 12 weeks (n=26), 20-24 weeks (n=21), and 36 weeks (n=17, 9 with one dose, and 8 with two doses; see Supplementary Material for full breakdown). Numbers decline over time as participants rebound. We only studied participants with viral suppression (plasma HIV RNA <200copies/ml) to avoid the confounding effects of plasma viraemia, which in itself stimulates an HIV-specific immune response. We used three antigen-specific assays: the Activation-Induced Marker (AIM) assay, the Cell Trace Violet (CTV) proliferation assay and the IFN-γ ELISpot assay – the first two with stimulation using HIV consensus B peptide pools (Gag, Env, Pol, or Nef) and the ELISpot using 15-mer overlapping full genome HIV peptides in a 2-dimensional megamatrix format consisting of 11-13 peptides/pool (**Supplementary Figure 1)**. In addition, we characterised unstimulated phenotypic and activation/exhaustion markers by flow cytometry after bNAb dosing, T cell receptor (TCR) repertoire changes, HLA Class I type, and single cell gene expression. These assays provided a comprehensive analysis of both the functional and proliferative capacity of T cells to respond to HIV antigens before and after bNAb dosing.

Although T cells recognise the full expressed HIV genome, responses to Gag-derived peptides have been linked with clinical control^16^. HIV Gag-specific IFN-γ ELISpot responses after bNAb dosing and ATI increased significantly from baseline (median 129 spot-forming units (sfu)/10^6^ PBMCs) through Week 12 (median 209 sfu/10^6^ PBMC, p=0.006), Week 20/24 (median 230 sfu/10^6^ PBMC; p=0.015), and sustained through Week 36 (median 250 sfu/10^6^ PBMCs P=0.034) (**Figure 1b**). This statistical result is not driven by those participants with lowest immune responses rebounding and dropping out, as the finding remains significant when only considering those represented at all timepoints (**Supplementary Figure 2a**). This is also demonstrated in Figure 1b-f, where open circles represent participants who sustained control for at least 36 weeks and closed circles show those who rebounded prior to this. To further validate *ex vivo* responses, we used the CTV proliferation assay to quantify HIV-specific memory T cell expansion. Proliferation of HIV Gag-specific CD8+ T cells significantly increased between Weeks 0 and 36 (median 7.5% vs 22.7%; p=0.006) and Weeks 0 and 12 for CD4+ T cells (1.9% vs 3.5%; p=0.01) (**Figure 1c-d; Supplementary Figure 2d**). T-cell activation and co-stimulatory molecules (CD25, CD134, CD137, CD69) were measured by the AIM assay. We found no statistical evidence of an increase in HIV Gag-specific AIM+ CD4+ and CD8+ T cells after bNAb dosing (**Figure 1e-f**). When Gag-specific ELISpot and proliferative responses were examined as a proportion of the total response across major HIV proteins (Gag, Pol, Env and Nef), there was a shift towards Gag-dominant responses 24 weeks after bNAb dosing compared to baseline (**Figure 1 g-i**). As a control, CMV responses were not impacted by HIV-specific bNAb therapy in any assay (**Figure 1j; Supplementary Figure 8**).

### Changes in immune responses to HIV Pol, Nef, Env after bNAbs

Immune responses to other HIV proteins (Pol, Nef, Env) were also analysed. There was a significant increase in HIV Pol-specific IFN-γ ELISpot responses and CTV-assessed CD4+ T cell proliferation from baseline to Week 12 (median 175 vs 233 sfu/10^6^ PBMC, p=0.013, and 2.72% vs 3.58%; p=0.041, respectively) (**Figure 2a-c**). In addition, the median frequency of AIM+ Pol-specific CD4+ T cells increased 36 weeks post-bNAbs, compared to baseline (0.20% vs 0.59% p=0.020) (**Figure 2d-e**). Immune responses to other HIV proteins were less marked, but with evidence for increased proliferation of HIV Nef-specific CD4+ and CD8+ T cells (0.07% vs 0.56%, p = 0.002 and 0.05% vs 0.65%, p = 0.004, respectively) between Weeks 0 and 36 (**Figure 2 f-g**). Full responses across all proteins are shown in **Supplementary Figure 2**.

**Figure 2.**
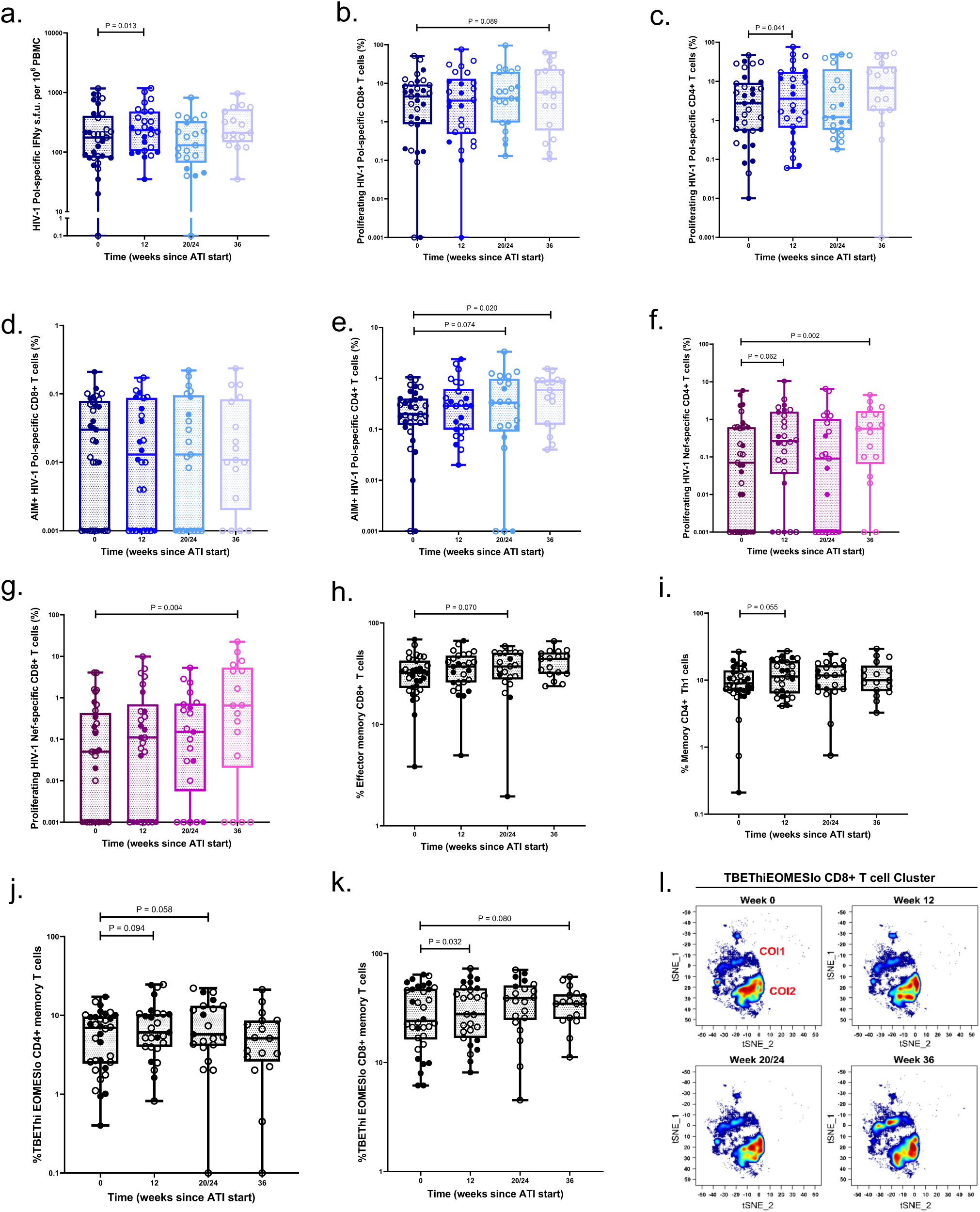
(a) HIV Pol-specific ELISpot, Proliferation ((b)CD8+ and (c) CD4+) and AIM ((d) CD8+ and (e) CD4+) assays 0, 12, 24 and 36 weeks after bNAb dosing and ATI in aviraemic participants. (f-g) HIV Nef-specific Proliferation (CD4+ and CD8+) assays 0, 12, 24 and 36 weeks after bNAb dosing and ATI in aviraemic participants. T cell phenotyping to show (h) CD8+ effector memory, (i) CD4+ Th1 (i) and TbetHiEomesLo (j) CD4+ and (k) CD8+ memory T cells. (l) Cytobank opt-SNE contour plots of betHiEomesLo CD8+ T cells. IFNy interferon gamma; s.f.u spot forming units; ATI Analytical treatment interruption.

### Phenotypic T cell changes after bNAbs

We next assessed general (i.e. not antigen-specific) T cell phenotypic differences pre- and post-bNAbs. There was no difference in the proportions of CD4+ or CD8+ T cells before and after bNAb dosing through Weeks 12 to 36, or between the frequencies of naïve, central memory (CM) and terminally differentiated effector memory (TEMRA) CD4+ and CD8+ T cells (**Supplementary Figure 3**). There was a trend for an increase in frequency of effector memory (EM) CD8+ T cells 24 weeks post-bNAbs and ATI (p=0.07) (**Figure 2h**) and Th1 CD4+ cells 12 weeks post-bNAbs (p=0.055) (**Figure 2i**). There was no evidence for an impact of bNAbs on the frequencies of exhaustion markers PD-1, TIGIT, TIM-3, and LAG-3 on memory CD4+ or CD8+ T cells (**Supplementary Figure 4**).

Greater expression of a Tbet^hi^Eomes^lo^ phenotype – associated with potent effector CD8+ T cell functions, and reduced T cell exhaustion - was observed in memory CD8+ T cells at Weeks 12 (p=0.03) post-bNAbs and ATI, with a trend for a sustained increase to Week 36 (P=0.08) (**Figure 2 j-k**). Interestingly, the Tbet^hi^Eomes^lo^CD8+ T cells were comprised of two sub-populations (labelled COI1 and COI2; **Figure 2l**), the former over-represented in controllers and marked by a decreased expression of CD45RA and TIGIT, potentially reflecting a population of a highly differentiated Tbet^hi^Eomes^lo^ cells with sustainable effector functions (**Supplementary Figure 4**).

### bNABs and ATI result in HIV-specific T cell clonal expansion

Six participants chosen to reflect a range of viral control (41-129 weeks suppression since last dose) were analysed further by single-cell transcriptomics, including TCR repertoire sequencing for five, according to sample availability, at baseline and Week 12 (details in Supplementary Material). Twelve weeks after first bNAb dose and ATI there was an increase in the number of expanded T cell clones, with a concentration in CD8+ cytotoxic T cell and MAIT cell populations (**Figure 3a-d**). Aligning TCR sequences with known specificities in the VDJ-db database (threshold of 85% identity in the CDR3 region of the beta chain) revealed an enrichment for Gag-specific CD8+ CTL clones with Pol and Nef representation, consistent with the findings in the functional assays (**Figure 3e**). Certain participants had remarkable clonal expansion after bNAbs, for example ‘Controller 4’ (**Figure 3f-g**), with multiple new medium-large clones, 344 of which matched known HIV-specific TCRs. Across all participants, Shannon diversity was higher at Week 12 than at baseline, consistent with the recruitment of novel, low-frequency clonotypes, that diversified the repertoire, despite concurrent appearance of the large clonal expansions.

**Figure 3.**
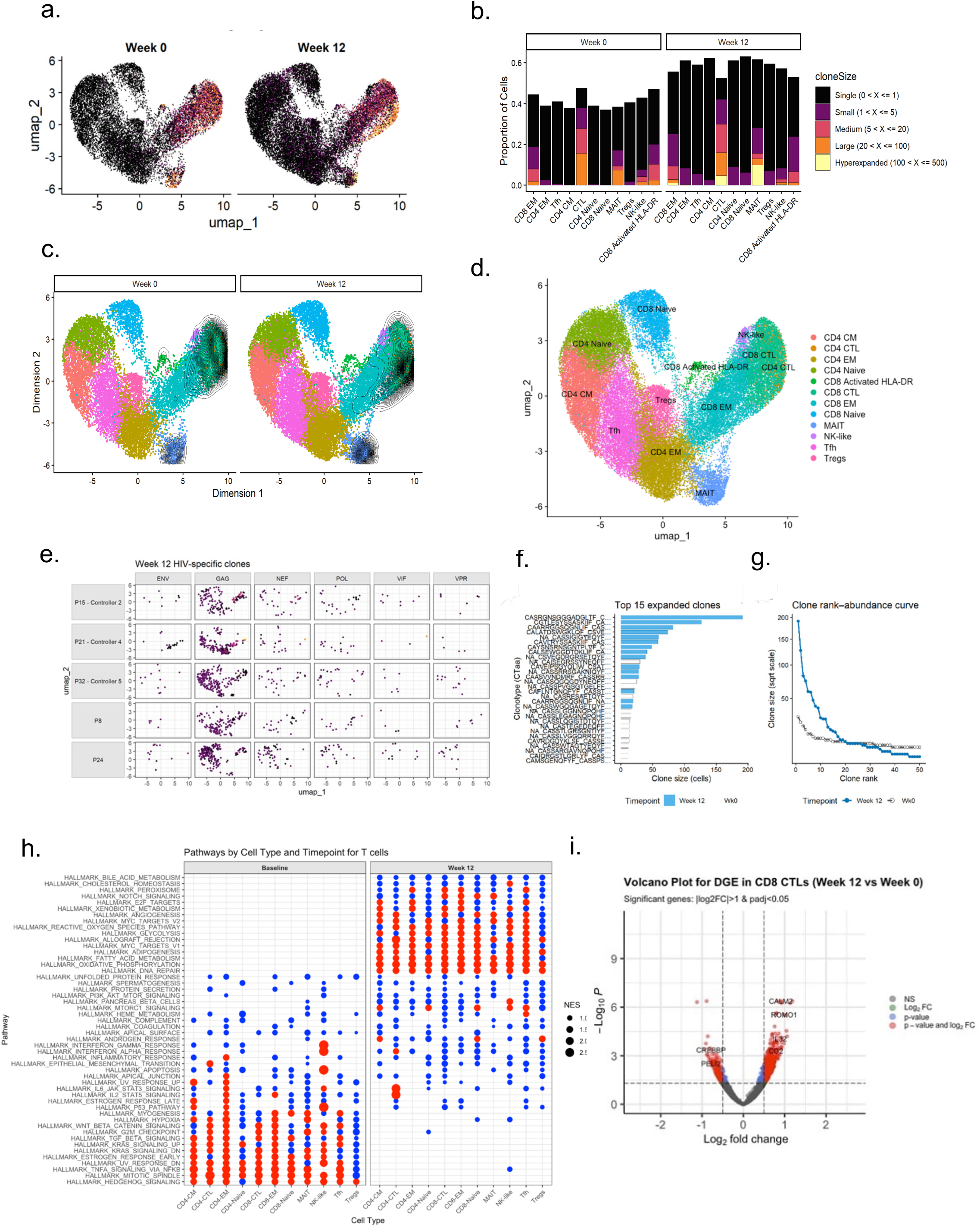
(a) UMAP embeddings of T cells coloured by expansion status at Week 0 and Week 12. (b) Distribution of clone-size classes across major T-cell subsets at baseline (Week 0) and 12 weeks post-bNAbs/ATI (Week 12). (c) Density contour plots showing cell types exhibiting expanded clones at both Week 0 and Week 12. (d) Un-stratified UMAP with annotated cell-types across groups. (e) Predicted HIV antigen specificity of TCRs (ENV, GAG, NEF, POL, VIF, VPR) projected onto UMAPs by participant, using VDJdb at 85% threshold identity. (f) Rank–abundance curve for the top 50 clonotypes expanded at Week 12 in Controller 4 (clones defined by unique paired TCR sequences). (g) Sizes of the top 15 expanded clonotypes in Controller 4 at Week 12 versus Week 0, labelled by CTaa (CDR3 amino-acid) sequences. (h) Gene set enrichment analysis (GSEA) of differential gene expression before and after bNAbs in 5 participants, and (i) volcano plot and of the same data.

Gene Set Enrichment Analysis (GSEA) (**Figure 3h)** was performed on the entire differentially expressed gene set (ranked by log2FoldChange × −log10 (padj + 1e-10), using the Hallmark pathway collection across all T cell subtypes. This analysis confirmed a transition after bNAbs and ATI from a stressed, inflammatory state (enriched for Hedgehog signalling, Mitotic Spindle, TNFa signalling via NFkB, and TGFb pathways) to a metabolically activated state (enriched for oxidative phosphorylation, fatty acid metabolism, MYC targets, and glycolysis). Although not reaching statistical significance, a trend towards enrichment of interferon-related pathways across all cell subtypes was observed following bNAb treatment.

Next, we looked in the cell subtypes where clonal expansion of TCRs was observed. A total of 747 genes were significantly differentially expressed in CTLs between participants treated with bNAbs 12 weeks post-ATI and baseline (|log2FoldChange| > 0.5, padj < 0.05) (**Figure 3i**). At Week 12, the predominant up-regulated genes were involved in metabolism and protein synthesis (NDUFB1, ATP5D, EIF2S1, ROMO1), reflecting high energy demands, active mitochondrial function, and enhanced translation. This profile is complemented by upregulation of immune and activation genes (CD2, IL32, GZMA, ICAM2). Overall, Week 12 CD8+ CTLs present a high-energy, active state. In contrast, at baseline, cells were enriched for chromatin regulators and transcription factors (KMT2D, EP300, CREBBP, ARID1A, PRDM1, RUNX3) as well as RNA processing genes (POLR2A, UPF1, ELL2, MEX3C), consistent with being less functionally active.

### Immune predictors of sustained virologic control after bNAbs and ATI

Of the 34 participants who received dual LS-bNAbs, three withdrew from the study early (<20 weeks) after the treatment intervention, and one died of a cause considered unrelated at Week 20. Loss of virologic control during ATI was defined as a sustained HIV plasma viral load (pVL) ≥1000 copies/ml for six weeks or two pVLs >100,000 one week apart. To investigate if HIV Gag-specific baseline immunological measurements were associated with time to viral rebound from ATI, we conducted an exploratory Cox proportional hazards analysis. To account for the different number and timing of bNAb doses, a time-dependent covariate for second-dose administration was added to the model. Due to the high dimensionality and potential multicollinearity among the immunological assays, LASSO regularisation was applied for variable selection.

For analysis of viral rebound, LASSO selected all five Gag-specific baseline T cell measures considered: Gag-specific CD4+ and CD8+ T cell proliferation, CD4+ and CD8+ AIM responses, and IFN-γ ELISpot responses. These variables were included in a multivariable time-dependent Cox proportional hazards model which showed that higher baseline Gag-specific CD8+ AIM responses were associated with a longer time to rebound (HR = 0.35; 95% CI: 0.18, 0.70; p = 0.003), while higher baseline Gag-specific IFN-γ ELISpot responses were associated with a shorter time to rebound (HR = 4.57; 95% CI: 2.02, 10.3; p <0.001) (**Figure 4a**). When only considering participants with no evidence of significant baseline bNAb-resistance associated mutations (RAMs)(n=28), there was evidence for CD8+ proliferation also being protective (HR = 0.33; 95% CI: 0.11–0.97; p = 0.043) (**Supplementary Figure 5).**

**Figure 4.**
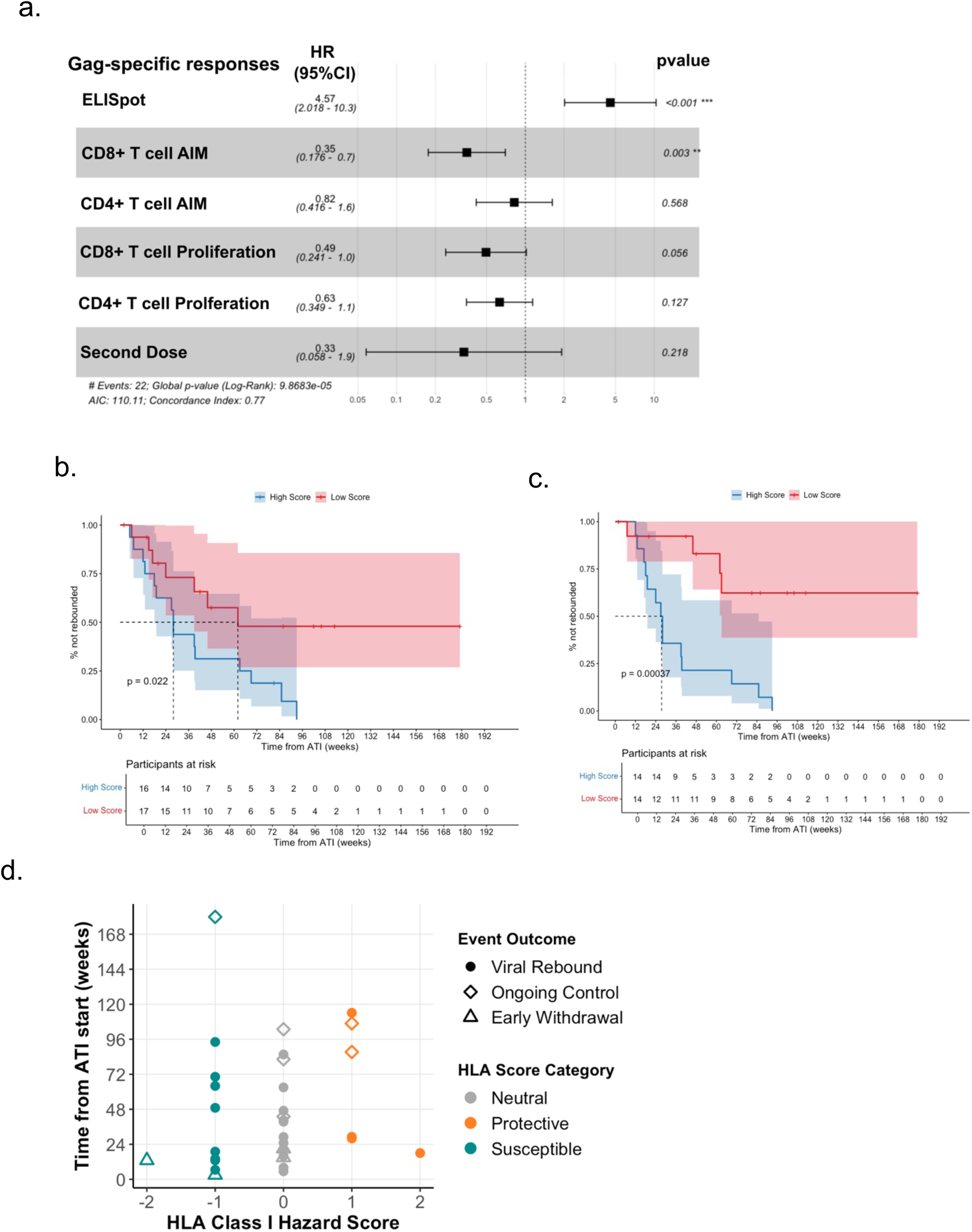
Multivariable Cox models using (a) baseline immunological data to predict time to rebound after ATI. Kaplan Meier survival based on immune score (IS) to predict viral rebound for (b) all, and (c) participants without baseline RAMs. Participants are segregated at the median IS. (d) Correlation plot showing between time to rebound and HLA Class I ‘Hazard Score’ including all participants.

To investigate if the increase in the magnitude of Gag-specific T cell immunity observed after bNAbs would predict control, we evaluated the association with time to viral rebound and the fold-change in responses between baseline, Week 12, and Week 24. As before, LASSO regularisation was used to select variables from the five Gag-specific immune measures to be included in a multivariable Cox model. Fold-change increases in CD4⁺ T-cell proliferation from Week 12 to Week 24 were associated with a reduced risk of viral rebound (HR = 0.52, 95% CI: 0.27–1.0, p = 0.049) (**Supplementary Figure 6a**). CD8⁺ T-cell proliferation and ELISpot responses were also selected by LASSO, but were not significantly associated with time to rebound. For changes from baseline to Week 24, LASSO selected fold-change of CD8⁺ T-cell AIM as associated with a lower risk of rebound, but this effect was not statistically significant in the Cox model (HR = 0.78, 95% CI: 0.48–1.25, p = 0.314) (**Supplementary Figure 6b**). No changes in immune responses at Week 12 from baseline were associated with time to rebound.

The contrasting impact of baseline Gag-specific CD8 AIM and ELISpot was demonstrated in a univariable survival analysis using a derived ‘immune score’ (IS) based on the effect estimates of the Cox model, as has been implemented for gene expression studies^17^. The hazard ratios obtained from the model were used as weighting coefficients to reflect the relative contribution of each immune parameter to outcome risk. The immune score for each participant was then calculated as: IS= (Cox Coefficient_ELISpot_ * ELISpot result) + (Cox Coefficient_CD8 AIM_ * CD8 AIM result). Participants were split into a high and low IS score groups based on the median. Kaplan Meier (KM) survival showed that a higher IS was significantly associated with faster rebound (P=0.022; HR=1.83, 95% CI: 1.29, 2.59; **Figure 4b**). When only considering participants without baseline sequences containing RAMs, CD8+ T cell proliferation was also identified as predictive parameter in a Cox model, and when included in the IS, the association was strengthened (KM p=0.0004; HR=2.43, 95% CI:1.57, 3.75; **Figure 4c**).

### HLA Class I and clinical outcomes after bNAb dosing

Certain HLA Class I alleles such as HLA B*5701 and B*2705 have been associated with clinical protection, either in the context of viral control, CD4 T cell decline or time to an AIDS diagnosis^18,19^. Other alleles such as B*3502 have been linked to more rapid disease progression^20^. To determine the association between HLA Class I and the response to bNAbs and ATI – and by proxy, evidence for T cell mediated control – we compared the time to viral rebound among participants with known HIV disease-protective or -susceptible alleles. Given the predominant White male demographic in RIO, only HLA Class I alleles previously shown to have the strongest association with protection or susceptibility to HIV progression in individuals with clade B viruses were considered^21^ (**Supplementary Figure 7a)**. Participant time to rebound from ATI start was analysed by creating a HLA Hazard Score (HS; protective (HS > 0), susceptible (HS < 0), and neutral (HS = 0); see Methods; **Figure 4d**) to account for the potential additive effects of alleles. We used a Cox proportional hazard model with a time-dependent covariate for second-dose administration. We found no evidence of an association between HLA Class I hazard score group and viral control, whether including all participants (**Supplementary Figure 7b**) or restricting to those without evidence of baseline bNAb RAMs (n = 31, **Supplementary Figure 7c)**. Additionally, when participants were grouped by a discrete HLA classification: protective (≥1 protective allele only), susceptible (≥1 susceptible allele only), mixed (both protective and susceptible alleles present), or neutral (neither present), there was no evidence of a difference in time to viral rebound across groups when compared to those with neutral HLA risk (**Supplementary Figure 7d**).

### Immunological Responses in Controllers

Seven participants (all of whom received two doses of bNAbs) sustained viral control without meeting the trial rebound criteria for over 96 weeks, with predicted bNAb levels below the presumed therapeutic threshold of 10ug/ml^15^ (**Figure 5a-b**). Three of these ‘Controllers’ (C1, C3, C4) had HLA Class I alleles associated with control (B*52:01 and B*57:01) and one with an allele associated with post-treatment control (C2; B*35:02). As we have previously reported that there were two post-treatment controllers (PTCs) in the placebo arm^15^, it is likely that a component of control seen in the intervention arm may be PTCs who also received bNAbs. **Figure 5c** shows the longitudinal immune responses for the different assays and bNAb concentrations across 36 weeks after bNAb dosing and ATI for the controllers. Although there is no consistent pattern of change over time, these seven controllers had median baseline Gag-specific CD8+ T cell proliferation (P=0.038; **Figure 5d**) and CD4+ AIM (P=0.048; **Figure 5e)** assay responses significantly greater than non-controllers. The median baseline ‘Immune Score’ of these seven participants was also significantly lower than the rest of the cohort (P=0.002) (**Figure 5f**).

**Figure 5.**
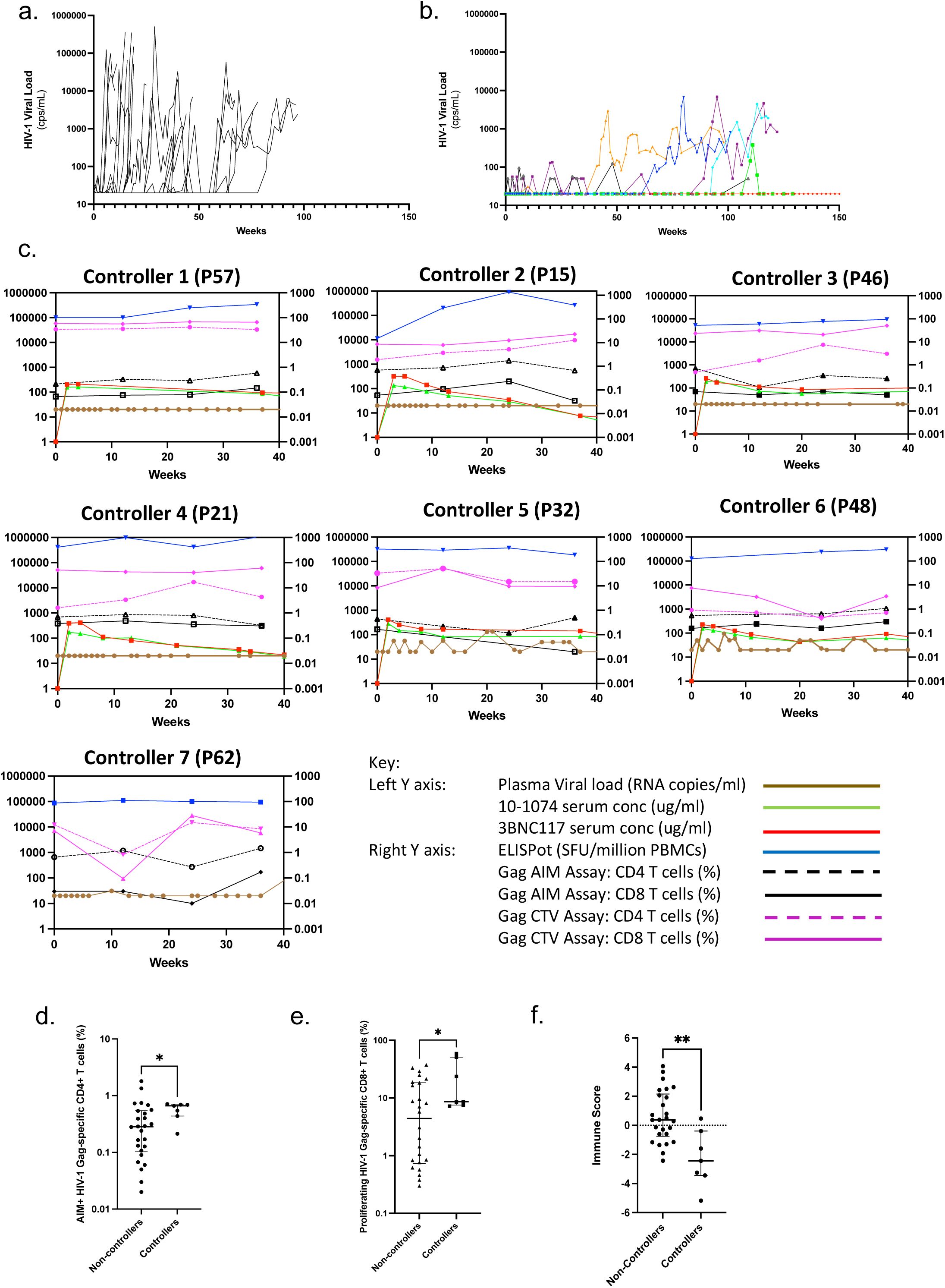
HIV plasma viral load plots for (a) RIO Arm A minus the seven participants controlling beyond Week 96, and (b) the seven controllers beyond 96 weeks. (c) Detailed viral load (brown), ELISpot (blue), CD4+ (hashed line magenta)and CD8+ proliferation (full line magenta), CD4+ (hashed line black) and CD8+ AIM data (full line black), 10-1074 serum concentations (ug/ml)(green) and 3BNC117 serum concentrations (ug/ml)(red) for the seven ‘controller’ participants for 36 weeks. Viral load (HIV RNA copies/ml)and bNAb concentration (ug/ml) values are represented on the Left Y axis; ELISpot (SFU/million PBMCS), proliferation (%) and AIM (%) data are presented on the Right Y axis. Non-parametric Mann-Whitney analyses of (d) CD8+ T cell proliferation, (e) CD4+ AIM responses and (f) the ‘Immune score’ from Figure 4c applied to the seven controllers vs non-controllers.

## Discussion

The mechanism and clinical importance of the proposed interaction between passively-administered therapeutic antibodies and host cell-mediated immunity is unclear^22^. Over 100 years ago, Smith postulated that protection from diptheria challenge in guinea pigs was enhanced by combining diptheria toxin and anti-toxin, compared to anti-toxin alone^23^ with the combination leading to ‘active immunity’. In cancer, immune complexes between anti-CD20 monoclonal antibodies and tumour antigens result in new anti-tumour T cell function in mouse models^24^. In Rhesus macaques, post-bNAb SHIV control is sustained by T cells, evidenced by rebound viraemia following infusion of anti-CD8 monoclonal antibodies^10^.

Further mechanistic insights are revealed by modification of the Fc region of antigen-specific monoclonals further enhancing this T cell vaccinal effect, as shown in animal models with Fc ‘GAALIE’ mutations in influenza^25^ and SARS-CoV-2^26^. However, despite these data and strong evidence for natural CD8 T cell-driven protection in ‘HIV elite controllers’ and genetic associations between HLA Class I and viral control^27^, the failure of T cell vaccines^28^, viral immune escape^29^ and non-reversible T cell exhaustion^30,31^ have questioned the enduring value of T cells in viral control and cure strategies.

To determine whether HIV-specific bNAbs induced a protective T cell vaccinal effect in the RIO trial, we assumed that two criteria needed to be met. Firstly, there should be new or enhanced HIV-specific immunity after bNAb dosing in participants who remained aviraemic. Secondly, there should be a statistically significant and meaningful association between this new or enhanced T cell immunity and clinical outcomes. We examined HIV-specific T-cell immunity, T cell phenotype, TCR repertoire diversity and single-cell resolution gene expression, HLA-associated risk and clinical outcomes in up to 34 people with HIV for 36 weeks after bNAb dosing and ATI.

Cellular assays revealed consistent and significant enhancement of baseline HIV-specific T-cell immunity post-bNAbs, primarily targeting viral structural proteins, detectable through the induction of interferon gamma (IFN-γ), cellular proliferation or activation markers. We observed enhanced IFN-γ producing HIV-specific T cells directed against the structural proteins Gag and Pol as well as increased proliferative and AIM responses, all in virally suppressed individuals following bNAb administration and ATI. As controls, we observed no changes in the frequency of CMV-specific T cell responses pre- and post-bNAbs and ATI by any assay, suggesting that the bNAb induced vaccinal effect was specific to HIV. Although not all assays to all peptides showed increases across all time-points, there was consistent evidence – particularly when stimulating with Gag or Pol proteins – of increased magnitude of responses, and a shift towards Gag-dominant responses, both of which would suggest enhanced T cell-mediated protection.

Phenotypic and functional T cell analyses showed a trend for an increased frequency of EM CD8+ T cells and T helper 1 cells post-bNAbs and ATI. The significantly higher expression of memory T cell differentiation markers Tbet^hi^Eomes^lo^ on CD4+ T cells and CD8+ T cells is also consistent with enhanced effector memory CD4+ T cell differentiation and functionality, not dissimilar to responses expected after an effective vaccine^32^. The increase in TCR repertoire clonality – in particular with the concentration in CD8+ CTL and MAIT cells – and the shift towards expressed genes associated with greater T cell functionality also indicate an enhancement of HIV-specific T cell immunity. Together, these data support the first premise that bNAbs with ATI are associated with new and enhanced HIV-specific T cell immunity.

Using ‘time to rebound’ from ATI as a marker of clinical efficacy, we found strong associations with clinical outcomes and baseline T cell immunity and – although to a lesser extent - with the change in responses after bNAb dosing and TI. It was the baseline responses that were most predictive and, after adjusting for other Gag-specific T cell measures, baseline measures of two T cell assays dominated clinical outcomes after ATI. Higher levels of HIV Gag-specific CD8+ AIM responses were protective (i.e longer duration of viral control after ATI), but Gag-specific ELISpot responses conferred harm – those participants with high baseline ELISpots were more likely to rebound early. Accordingly, the group predicted to control for longest after bNAbs were those with high CD8 AIM and low ELISpots, as was strongly indicated in the ‘Immune Score’ survival analysis. When focusing on those participants without baseline RAMs, CD8+ T cell proliferation also predicted outcomes. This is consistent with findings in the eClear study, which associated sensitivity to bNAbs with stronger T cell immune responses, hypothesised to be due to more efficient immune complex formation and presentation on antigen-presenting cells ^4,7^.

The ELISpot result was unexpected. It has been shown that HIV-specific immunity declines on fully suppressive ART^33,34^, however there is much inter-patient variability. Whether high ELISpot responses on virally-suppressive ART reflect greater viral antigen exposure from a more transcriptionally-active reservoir with a greater likelihood of early rebound is worth exploring, and would argue for its role in pre-ATI screening. The only evidence for an increase in T cell immunity after bNAbs being protective was the change in Gag-specific CD4+ T cell proliferative responses from Week 12 to 24. This result demonstrates that changes that occur after bNAbs and ATI may also drive outcomes. Although this therefore meets our second criteria of the induced response impacting clinical outcomes and confirming a T cell ‘vaccinal effect’, our data strongly indicate that the baseline T cell responses are the best predictors of outcome. However, a separate open-label study of 10 participants showed no association between baseline HIV-specific responses and post-bNAb control^11^. Interestingly, this study was a mixture of early and late infection whereas RIO was just early, and one explanation for this discrepancy could be the accrual of T cell dysfunction and exhaustion over time in chronic HIV.

Focussing on the seven participants who sustained the longest control (not meeting the trial endpoint within 96 weeks of ATI) there was again strong evidence of baseline immunity predicting clinical outcomes. As these participants required ART during their initial primary infection but were subsequently able to remain off therapy for two years after bNAbs, this suggests a fundamental switch in the host-virus dynamic. The ontogeny of these immune responses could be, at least, two-fold. T cells are remarkably sensitive measures of antigen exposure, as used clinically for latent TB diagnosis^35^. As such, one may need to distinguish between more efficient antigen presentation by bNAb-HIV immune complexes and the enhanced antigen load from viral reactivation in the ATI itself. In the placebo arms of ATI trials, rapid resurgence of viraemia is seen within a few weeks^15,36^. The same viral antigen reactivation would be expected in RIO on stopping ART, however bNAbs prevented detectable viraemia. Whether HIV-specific T cell memory is stimulated through direct antigen presentation from infected cells on HLA Class I or through immune complexes interacting with antigen presenting cells requires further analysis. Our cytometry panels did not include markers for ‘stemness’, such as TCF-1, and it is interesting to note recent publications which suggest that post-bNAb control may be the result of boosting the proliferative potential of these pre-existing, high quality stem-like T cells^8,11^. However, together our findings support the role of HIV-specific T cell proliferative potential being associated with control after bNAb dosing

The study has certain limitations. Although there is clear evidence for the virally-suppressive impact of bNAbs, the interpretation of serum levels and PK analysis is complicated by the lack of a defined clinically-relevant concentration above which they are effective. In addition, serum bNAb concentrations associated with neutralisation may be different to those associated with Fc functions or immune complex formation, and this may also vary across different compartments such as lymph nodes, CSF and serum. We have previously shown that the median serum bNAb concentrations at the time of viral rebound in RIO for 10-1074LS and 3BNC117LS were 40.6 µg/mL (range 0.6 – 149 μg/mL) and 52.4 µg/mL (range 0.4 – 231.4 μg/mL), respectively^15^. All six participants with available bNAb measurements who remained off ART at 96 weeks had measured bNAb rebound concentrations below 10 μg/ml. Simulations of the time for LS-bNAb concentrations to reach 10μg/mL after two doses given 21 weeks apart was 48.6 weeks (90% PI 37.1 – 59.0) and 47.8 weeks (90% PI 38.6 – 57.1) after the second dose, for 10-1074LS and 3BNC117LS, respectively^15^. The other complexity for the analysis is the heterogeneity of bNAb dosing, with half the cohort receiving one dose and the other half receiving two doses, between 21 and 50 weeks after randomisation. To account for this we used a time-dependent Cox proportional hazards model which accounted for those receiving one or two doses. Although no model in this scenario is gold standard, other approaches used (eg measuring rebound times from ‘last dose’ as well as from ‘initial ATI’ with censoring at the second dose timepoint) produced consistent results.

In conclusion, these data demonstrate a clinically protective post-bNAb T cell vaccinal effect. The divergent associations between measures of baseline T cell immunity and clinical outcomes may reflect that not just the size, but also the volatility, of the HIV latent reservoir is a key consideration. Many questions remain unanswered, such as the role of innate immunity and autologous neutralisation after bNAb dosing, work which is on-going^37^. However, these data show that bNAb dosing facilitates HIV-specific T-cell responses with the quality to control viral replication off ART, and suggest that long-term ART-free HIV remission may be achievable with the correct combination of interventions such as bNAbs and an effective host immune response.

## Methods

### Study design and participants

RIO was a prospective multi-centre phase II double-blinded randomised placebo-controlled trial performed according to the principles of the Declaration of Helsinki (NCT04319367). Ethical approval to undertake the research was obtained from UK Health Research Authority, and Research Ethics Committee (REC) approval was granted by the London-Westminster REC. UK Research Ethics Committee reference: 19/LO/1669 (11 Sep 2019); EudraCT: 2019-002129-31 (12 Dec 2019); EU CTR: 2024-514564-13-00 (2 Jan 2025). Written informed consent was obtained from all participants prior to inclusion in the study. Participants were screened for predicted 10-1074 sensitivity through sequencing single proviral Env amplicons and those with RAMs present in >15% of sequences were excluded from the trial. Sixty-eight participants on ART since three months of primary HIV were enrolled between 2021 and 2024 in the UK and Denmark. Participants were randomised 1:1 to receive (Arm A) two LS-bNAbs (3BNC117 and 10-1074) or (Arm B) placebo (normal saline) and an ATI. The demographics and clinical characteristics of the patients are shown in **Table 1 and Supplementary Table 1**. Of the 34 patients in Arm A who underwent treatment intervention, three patients withdrew from the study and one patient died from cardiovascular events which were not felt to be associated with bNAbs or ATI. Blood samples were collected pre-bNAb administration and after Weeks 12, 20/24, and 36. Only aviraemic individuals in Arm A (HIV RNA viral load < 200 copies/mL) were considered for this analysis.

### Analysis of Time to Rebound

Viral rebound was determined by an independent Endpoint Adjudication Committee as the first detectable viral load resulting in either six weekly consecutive readings >1000 copies/ml or two readings >100,000 one week apart. For this immunological study, if there was any clinical variation from these simple outcomes, a conservative approach was taken to allocate the date of rebound to avoid any risk of over-estimating post-bNAb control.

### Assessment of HIV-specific T-cell immunity

#### Ex vivo IFN-γ ELISpot

Cryopreserved PBMC were used to perform ELISpot assays. 50,000 PBMC/well were stimulated using 15-mer overlapping HIV peptides (Gag, Pol, Env, Nef) in a megamatrix consisting of 11-13 peptides/pool at 2 µg/mL per peptide (**Supplementary Figure 1**). CMV pp65 (2 µg/mL Miltenyi Biotec, Germany), FECT (Influenza, EBV, CMV, Tetanus) and phytohaemagglutinin-L (Sigma-Aldrich) were used as controls. After overnight incubation addition of streptavidin-alkaline phosphatase and BCIP-NBT Plus chromogenic substrate (Mabtech AB) was followed by analysis using ImmunoSpot (CTL ELISpot reader, Ohio, USA). Positive wells were reported as spot forming units (sfu) per million PBMC after subtracting background. Immune responses greater than the mean plus 2 SD of unstimulated wells were counted as positive. Total OLP responses for each protein were summed to provide the protein-specific response.

### Cell proliferation assay

For the T cell proliferation assay cryopreserved PBMCs were stained with CellTrace Violet (CTV, Life Technologies) and stimulated with pools of HIV peptide covering the Gag, Pol, Env, and Nef proteins (1 μg/mL). DMSO was used as a negative control. CMV pp65 peptide pools and PHA-L (5ug/ml) were used as positive controls. After seven days, outputs were read using a MACSQuant x10 (Miltenyi Biotec, Germany) with data analysed using FlowJo version 10. A 1% cut-off was used to define a positive response.

### AIM assay

The AIM assay was carried out using cryopreserved participant PBMCs using stimulation with HIV subtype B peptide pools (Gag, Pol, Env, or Nef; 2 µg/ml). CMV pp65 peptide pools PHA-L and DMSO were included as controls. Antibody panels are described in Supplementary Table 2. Data were acquired using a BD LSR II Flow Cytometer and analysed using FlowJo software. For attribution of AIM+ve status, CD4 T cells that were positive for both CD25 and one of CD134, CD137, or CD69 were included. Attribution for CD8 T cells was a sum of four gates comprising two or more of any of CD25, CD137 or CD69.

### Immune phenotyping

Immune phenotyping was undertaken with cryopreserved PBMCs using a combination of cell surface and intracellular antibodies (Supplementary Table 2), and acquired using a BD LSRII cytometer, FACSDiva software and analysis with FlowJo version 10.

### Gene Expression and T cell repertoire analysis

Single-cell RNA sequencing was performed to profile transcriptomic changes at the cellular level in six participants between two timepoints, baseline and 12 weeks post bNAb dose. PBMCs were processed using the 10x Genomics Chromium 5’ Next GEM v2 platform according to the manufacturer’s protocol. Briefly, viable single cells were encapsulated into Gel Bead-In-Emulsions (GEMs), and barcoded cDNA libraries were generated using reverse transcription and amplification. The libraries were sequenced using Illumina NovaSeq X Plus 150 PE, with a depth of 20000 rads per cell per sample in the library. The data were demultiplexed and aligned on the human transcriptome using CellRanger v8 on cloud.10xgenomics.com. Individual seurat objects were created using the aligned reads from each sample and the corresponding metadata. Low-quality cells with fewer than 200 genes were removed and mitochondrial and ribosomal genes (>10%) were regressed out and the most informative genes were identified based on their variance across cells. Principal component analysis (PCA) was performed to reduce dimensionality of data and identify the most significant source of variation. The appropriate number of principal components (PC) then the nearest neighbors were found for each cell based on the PC selected. Cell clusters were annotated based on canonical marker genes and differential gene expression and pathway analysis was performed downstream, using seurat functions. To enable sample-level differential expression analysis using DESeq2 (Love et al., 2014), pseudobulk profiles were generated by summing raw counts across all cells belonging to the same participant, time point, and cell type. Pathway analysis was performed using fgsea and the hallmark collection of pathways.

TCR (V(D)J) sequencing libraries were generated in parallel using the 10x Genomics Chromium Single Cell 5′ V(D)J kit. V(D)J libraries were generated on the 10x Genomics Chromium platform and processed with Cell Ranger to obtain V(D)J outputs, specifically per-contig annotations, clonotypes and consensus annotations. V(D)J contig tables for each sample were parsed with scRepertoire (combineTCR) to define clonotypes; barcodes were harmonised to allow dual TCR and gene-expression analysis using Seurat. Clonal expansion and diversity were summarised with scRepertoire and results visualised on UMAPs. Where indicated, putative antigen specificity was annotated by approximate matching of CDR3 sequences to VDJdb (downloaded 25.09.25) using Levenshtein distance (R stringdist), and per-sample hits were reported at 85% identity threshold in the beta chain CDR3 region.

### BNAb pharmacokinetics analysis

Serial serum bNAb concentrations were measured by a validated anti-idiotype ELISA. Pharmacokinetics analyses of serum bNAb measurements were modelled using a two-compartment model with Monolix^TM^ software using the Stochastic Approximation for Model Building Algorithm. The following covariates were tested: Age, Log(Weight), HIV-1 clade, predicted baseline bNAb resistance mutations, and number of doses. 90% prediction intervals (PI) were calculated using RsmLx in R (1000 bootstrap replicates). Further details previously published^15^.

### HLA typing and Analysis

DNA from granulocytes was extracted (Gentra Puregene Blood Kit, QIAGEN, USA) and underwent HLA Class I and II typing via sequence specific amplication (SSP). The following alleles were classified as protective: A25:01, B13:02, B27:05, B52:01, and B57:01; and as susceptible: B07:02, B08:01, and B35 (B35:01, B35:02, and B35:03). An HLA hazard score (HS) was calculated for each participant by summing their six alleles, assigning a +1, −1, or 0 to alleles classified as protective, susceptible, or neutral, respectively, to reflect a potential additive effect.

### Time-dependent Cox proportional hazards model

To assess whether baseline immune responses or HLA Class I were associated with the timing of rebound, we used a Cox proportional hazards model that incorporated the administration of a second bNAb dose as a time-dependent variable. This approach allowed participants to contribute person-time both before and after receiving the second dose, thereby accounting for changes in risk that occur once the second dose was given. Participants without a recorded second dose were assigned an effective second-dose time of infinity, indicating that a second dosing did not occur during follow-up. A time-to-event data structure was created to allow the inclusion of time-dependent covariates. The follow-up time was defined by the interval between study entry and either relapse or censoring with the appropriate event status. The second_dose variable was modeled as a binary, time-dependent covariate that changed from 0 to 1 at the time of second-dose administration.

### Immune Score

The ‘immune score’ (IS) was created using the outcome of the Cox model to summarize the combined effect of significant immune predictors on clinical outcome. The hazard ratios (Cox coefficients) from the model were used as weighting factors to represent the relative contribution of each statistically significant immune parameter to outcome risk. For each participant, the immune score was calculated as:

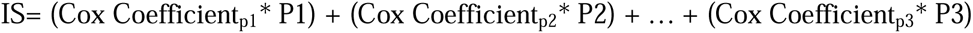

where P_i_ denotes the value of the ith immune parameter.

### Statistical analysis

GraphPad Prism 10 (San Diego, CA, USA) and R were was used for basic statistical analyses. Patient demographics and clinical characteristics were represented as median (IQR) and n (%), respectively. Statistical comparisons were made using a two-tailed non-parametric paired *t* test (Wilcoxon signed ranked test). Box plots were used to represent median (horizontal line), 25th and 75th percentiles (boxes) and minimum and maximum values (whiskers). Immune assay data were processed prior to time-dependent Cox proportional hazards modelling. The data were log-transformed to reduce skewness and scaled to facilitate comparison across assays. Log-fold changes were computed relative to baseline using the smallest non-zero value within each assay as the denominator to prevent division with zero values. The tmerge function function was used on R to create a time-to-event dataset that recorded each participant’s follow-up period and incorporated the timing of the second dose as a time-dependent variable.

## Data Availability

All data produced in the present study are available upon reasonable request to the authors

## Acknowledgements

We gratefully acknowledge all participants and the clinical staff at each site involved in the RIO Trial. The complete list of staff is available in the supplementary materials. The RIO Trial was funded by The Gates Foundation (OPP1210792). JF acknowledges support from the NIHR Oxford Biomedical Research Centre and the Martin Delaney REACH Collaboratory. S. Fidler, and H.B., and the Imperial Clinical Trials Unit acknowledge support from the NIHR Imperial Biomedical Research Centre (BRC). MCN and MC acknowledge support from the Stavros Niarkos Foundation and Robert Wennett. MCN is a Howard Hughes Medical Institute (HHMI) Investigator. Infrastructure support for this research was provided by the NIHR Imperial Biomedical Research Centre (BRC) and the NIHR Imperial Clinical Research Facility (CRF). MJL received funding from the Medical Research Council UK for a Clinical Research Training Fellowship (MR/W024454/1). KES and GDT acknowledge funding by the Gates Foundation (INV-036842) and the NIH Center for AIDS Research (P30AI064518). We acknowledge the NIH HIV Reagent Programme and BEI Resources Repository for the provision of HIV peptides. The remaining authors do not have any competing interests to declare. The funders were not involved in the study design, data collection, analysis, or interpretation of data, the writing of this article, or the decision to submit it for publication.

## Author contributions

See Supplementary Material

