## Supplementary Material for "T cell immunity predicts clinical outcomes on stopping antiretroviral treatment after HIV-specific broadly neutralising antibody therapy"

### RIO TRIAL INVESTIGATORS

**Chief Investigator:** Sarah Fidler; **Protocol Co-Chair:** John Frater
**Co-Investigators:** Michel Nussenzweig, Marina Caskey

**Trial Investigators:** Sarah Fidler, John Frater, Graham Taylor, Michel Nussenzweig, Marina Caskey, Saye Khoo, Ming Lee, Simon Collins, Julie Fox, Amanda Clarke, Sabine Kinloch-de Loes, Sarah Pett, Kyle Ring, Chloe Orkin, Marta Boffito, Gary Whitlock, Rebecca Sutherland, Alison Uriel, Marcelino Molina, Louise Terry, Ole Schmeltz Søgaard, Jesper Damsgaard Gunst, Henrik Nielsen, Jesal Gohil, Tamara Elliott, Hanna Box, Stephen Fletcher, Louise-Rae Cherrill, Emanuela Falaschetti, Daphne Babalis, Christina Prechtl, Najwa Soussi, Milaana Jacob, Ambreen Ashraf, Toby Prevost, Nicholas Johnson, Mariam Habib, Marta Gielniewska, Jodi Meyerowitz, Mary Cross, Verity Leeson, Amnah Mirza, Jonathan Dao, Eloise Britten, Vivienne Okona-Mensah, Amanda Bravery, Francesco Lala, Nayan Das, Smita Das, Lee Barker, Maathini Balachandran, Katie Topping, Jacquie Ujetz, Ishrat Jahan, Andrew Lovell, Maryam Khan, Helen Brown, Nicola Robinson, Mohammed Altaf, Penny Zacharopoulou, Timothy Tipoe, Ane Ogbe, Matthew Pace, Marcilio Fumagalli, Anna Kaczynska, Cintia Bittar

**Community Representatives:** Simon Collins, Ben Cromarty, Jo Josh, Roy Trevelion

**Trial Steering Committee (TSC):** Frank Post (Independent Chair), Caroline Sabin, Clifford Leen, Jaime Vera, Dimitra Peppa, Ben Cromarty

**Independent Data Monitoring Committee (IDMC):** Abdel Babiker (Independent Chair), Jane Anderson, Andrew Lever, Jo Josh, Roy Trevelion (previous member)

**Endpoint Adjudication Committee (EAC):** Linos Vandekerckhove (Independent Chair), Beatriz Mothe Pujadas, Casper Rokx, Ole Schmeltz Søgaard (previous Chair)

**Investigators and Staff at Participating Sites:**

**Sarah Fidler**, Ming Lee, Marcelino Molina, Jesal Gohil, Tamara Elliott, Euan Sutherland, Wilbert Ayap, Claire Petersen, Clive Matthews, Ian McGuinness, Charlotte Blake, Sophie Taylor, Romina Tajik, Rebecca Hall, Zoe Wiggins. **Sabine Kinloch-de Loes**, Jonathan Edwards, Thomas Allan, Katie Spears, Thomas Fernandez, Jia Bo He, Abigail Tobin, Fiona Burns, Megan Bailey, Mike Young, Pedro Simoes, Nnenna Ngwu, Margaret Johnson, Qayo Egeh. **Julie Fox**, Louise Terry, Piyumika Godakandaarachchi, Kathy Arbis, Alice McKain, Jacqueline O’Connell, Anele Waters, Julianne Lwanga, Harry Coleman, Isabel Laszczak, Rebekah Roberts, Mariusz Racz. **Chloe Orkin**, Kyle Ring, John Thornhill, James Hand, Helena Miras, Hafiza Rahman, Isabelle Whelan, Harriet Le Voir, Hamzah Farooq, Louise Whitton. **Sarah Pett**, Sandra Coombes, Deirdre Sally, Manik Kohli, Vasanth Naidu, Alejandro Arenas-Pinto, Felicity Aiano, Gosala Gopalakrishnan, Florence Bascombe, Joe Phillips, Ebunoluwa Taiwo, Claudia Benatar, Irfaan Maan, Marzia Fiorino, Charlotte Woodward, Jose Paredes Sosa, Rhiannon Owen, Claire Mullender, Nicola Stewart, Emmi Suonpera. **Marta Boffito,** Gary Whitlock, , Rizamay Balista, Serge Fedele, Alfredo Soler Carracedo, Betsabe Rodriguez Mateos, Rosalie Housman, Roya Movahedi, Ruth Byrne, Hyun Jin Kim, Freya Toyne, Ana Milinkovic, Julie Logan, Jad Salha, Clovis Salinos, Reda Yassein, Rosa Diaz Echeverria, Patrizia Simonato, Cherry Colcol, Veronica Canuto, Krestine Electito. **Amanda Clarke,** Kiersten Simmons, Tanya Adams, Mikaela Vinick, Lisa Barbour, Caroline Cable, Andrea Terlingo, Claire Norcross, Sophie Ross, Sonia Raffe, Sophie Ross, Fionnuala Finnerty, Vittorio Trevitt, Bryony Broster, Leigh Greenland, Celia Richardson, Andrew Bexley, Sarah Kirk, Collins Iwuji, Justina Strikaite, Goabaone Diteko, Donards Kim Tanedo, Marion Campbell. **Alison Uriel,** Denise Kadiu, Milisha Gihan, Gabriella Lindergard, Claire Fox, Chloe Lord, Irvine Mangawa, Pabalelo Pule, Pamela Hackney, Jacinta Guerin, Phoebe Shellman, Serah Titus, Raiza Hussain, Sam Hey, Bini George, Yu Tang, Kevin Kuriakose, Karen Talbot, Lisa Southon, Thomas Lamb, Fahd Niaz, Alice Hendy, Jan Flaherty. **Lisa Hamzah,** Aline Lopes Fernandes, Joana Teixeira, Hong Ju, Maria Belini, Samantha Taylor, Claire Gilmartin, Alkida Bucaj, Bojana Dragovic. **Rebecca Sutherland,** David Dockrell, Jurgen Haas, Susie Ferguson, Amy Shepherd, Louise Sharp, Jacqueline Henderson, Karla Berry, Anna Gordon, Alexandria Chung, Penelope Saverton, Jennifer Marshall, Anne Saunderson. **Ole Schmeltz Søgaard,** Jesper Damsgaard Gunst, Ane Søndergaard, Ida Soelberg Høj, Yordanos Yehdego, Sandra Schieber, Lene Svinth Jøhnke, Marie Juul Eli. **Henrik Nielsen,** Maria Ruwald Juhl, Kristine Toft Petersen, Rikke Thisted. **Paola Cicconi,** Vanessa Fenech, Juliana Umunna, Musaiwale Kamfose, Charlotte Wells, Mohammad Iqbal, Mehreen Datoo, Vaitehi Nageshwaran. **Karen Mosley,** Lisa Hurley, John Bervin Galang, Susanne Fagerbrink, Marcus Frederick, Christos Karathanasis, Victoria Tsui, James Fletcher, Tom Cole, Sofia Coelho, Oluwatoyin Ayanjoke, Gifty Teibowei, Ernesto Angustia, Guillermo Batan, Konstantina Pastou, Natalie Man, Aneta Gupta, Orla Mulrooney, Cyndi Cruzata, Alanood Alshamari, Zoe Gardener, Theo Farah, Peter Talbot, Maniola Tanaka, Abiola Ogunleye, Joanna Schronce, Peace Usigbe, David Owen, Janahi Visakan, Maria Renguenge, Asha Vikraman, Bridget Oduro, Roisin O’Sullivan, Andrew Ravendren, Daisy Metcalf, Nicola Lawlor, Suryapriya Thovaray Balan, Sam Jawaid, Cecilia Njenga, Nana-Marie Lemm, Mumsy Mahange, Kim Sorley, Shelley Page, Relebogile Mawasha, Derecia Adelakun-Powlette, Tamunoibim Anidima, Sukumuran Ajithkumar.

**Pharmacokinetic (PK) Assay laboratory**.

**Kelly E. Seaton**, Shad Mosher, Alex Carnacchi, Sheetal Sawant, **Georgia D. Tomaras**

**Program management**

Hongmei Gao, Kelli Greene

**Anti-Drug Antibodies Assays**

Margaret E. Ackerman, Joshua A. Weiner

MANUSCRIPT CONTRIBUTIONS TABLE **(CRediT taxonomy; https://credit.niso.org/)**

| **Author** | **Number** | **Conceptualization** | **Data curation** | **Formal analysis** | **Funding acquisition** | **Investigation** | **Methodology** | **Administration** | **Resources** | **Software** | **Supervision** | **Validation** | **Visualization** | **Writing –draft** | **Writing – review** |
| --- | --- | --- | --- | --- | --- | --- | --- | --- | --- | --- | --- | --- | --- | --- | --- |
| **Mohammed Altaf** | 1* |  | **X** | **X** |  | **X** | **X** |  |  |  | **X** | **X** | **X** | **X** | **X** |
| **Carla Nel** | 1* |  | **X** | **X** |  | **X** | **X** |  |  |  |  | **X** | **X** | **X** | **X** |
| **Timothy Tipoe** | 1* |  | **X** | **X** |  | **X** | **X** |  |  |  |  | **X** | **X** | **X** | **X** |
| **Julia Edgar** | 1* |  | **X** | **X** |  | **X** | **X** |  |  |  |  | **X** | **X** | **X** | **X** |
| **Panagiota Zacharopoulou** | 1* |  | **X** | **X** |  | **X** | **X** |  |  |  |  | **X** | **X** | **X** | **X** |
| **Devinder Srai** | 6 |  | **X** | **X** |  | **X** | **X** |  |  |  |  | **X** | **X** | **X** | **X** |
| **Chanice Knight** | 7 |  | **X** | **X** |  | **X** | **X** |  |  |  |  | **X** | **X** | **X** | **X** |
| **Ming Lee** | 8 |  | **X** | **X** |  | **X** | **X** |  |  |  |  | **X** | **X** | **X** | **X** |
| **Louise-Rae Cherrill** | 9 |  | **X** | **X** |  |  | **X** |  |  |  |  |  |  |  | **X** |
| **Emanuela Falaschetti** | 10 |  | **X** | **X** |  |  | **X** |  |  |  |  |  |  |  | **X** |
| **Ane Ogbe** | 11 |  | **X** | **X** |  | **X** | **X** |  |  |  |  | **X** | **X** | **X** | **X** |
| **Stephen Fletcher** | 12 |  | **X** |  |  |  |  | **X** |  |  |  |  |  |  | **X** |
| **Hanna Box** | 13 |  | **X** |  |  |  |  | **X** |  |  |  |  |  |  | **X** |
| **Tsmara Elliott** | 14 |  | **X** |  |  | **X** | **X** | **X** |  |  |  |  |  |  | **X** |
| **Sabine Kinloch** | 15 |  | **X** |  |  | **X** | **X** | **X** |  |  | **X** |  |  |  | **X** |
| **Julie Fox** | 16 |  | **X** |  |  | **X** | **X** | **X** |  |  | **X** |  |  |  | **X** |
| **Amanda Clarke** | 17 |  | **X** |  |  | **X** | **X** | **X** |  |  | **X** |  |  |  | **X** |
| **Sarah Pett** | 18 |  | **X** |  |  | **X** | **X** | **X** |  |  | **X** |  |  |  | **X** |
| **Simon Collins** | 19 | **X** |  |  |  |  |  | **X** |  |  |  |  |  |  | **X** |
| **Maathini Balachandran** | 20 |  |  |  |  |  |  | **X** | **X** |  |  |  |  |  | **X** |
| **Katie Topping** | 21 |  |  |  |  |  |  | **X** | **X** |  |  |  |  |  | **X** |
| **Louise Terry** | 22 |  | **X** |  |  | **X** | **X** | **X** |  |  | **X** |  |  |  | **X** |
| **Kelly Seaton** | 23 |  | **X** | **X** | **X** | **X** | **X** | **X** |  |  |  | **X** |  |  | **X** |
| **Georgia Tomaras** | 24 |  | **X** | **X** | **X** | **X** | **X** | **X** |  |  |  | **X** |  |  | **X** |
| **Alison Uriel** | 25 |  | **X** |  |  | **X** | **X** | **X** |  |  | **X** |  |  |  | **X** |
| **Chloe Orkin** | 26 |  | **X** |  |  | **X** | **X** | **X** |  |  | **X** |  |  |  | **X** |
| **Kyle Ring** | 27 |  | **X** |  |  | **X** | **X** | **X** |  |  | **X** |  |  |  | **X** |
| **Gary Whitlock** | 28 |  | **X** |  |  | **X** | **X** | **X** |  |  | **X** |  |  |  | **X** |
| **Marta Boffito** | 29 |  | **X** |  |  | **X** | **X** | **X** |  |  | **X** |  |  |  | **X** |
| **Rebecca Sutherland** | 30 |  | **X** |  |  | **X** | **X** | **X** |  |  | **X** |  |  |  | **X** |
| **Ole Sogaard** | 31 |  | **X** |  |  | **X** | **X** | **X** |  |  | **X** |  |  |  | **X** |
| **Jesper Gunst** | 32 |  | **X** |  |  | **X** | **X** | **X** |  |  | **X** |  |  |  | **X** |
| **Helen Brown** | 33 |  | **X** | **X** |  | **X** | **X** |  |  |  | **X** |  | **X** |  | **X** |
| **Nicola Robinson** | 34 |  | **X** | **X** |  | **X** | **X** |  |  |  | **X** |  | **X** |  | **X** |
| **Gabriella LIndegard** | 35 |  | **X** | **X** |  | **X** | **X** |  |  |  | **X** |  | **X** |  | **X** |
| **Philip Goulder** | 36 |  |  | **X** |  | **X** | **X** |  |  |  | **X** |  |  |  | **X** |
| **Graham Taylor** | 37 | **X** | **X** |  |  | **X** | **X** | **X** |  |  |  |  |  |  | **X** |
| **Marina Caskey** | 38* | **X** | **X** | **X** | **X** | **X** | **X** | **X** | **X** |  | **X** |  |  | **X** | **X** |
| **Michel Nussenzweig** | 38* | **X** | **X** | **X** | **X** | **X** | **X** | **X** | **X** |  | **X** |  |  | **X** | **X** |
| **Sarah Fidler** | 38* | **X** | **X** | **X** | **X** | **X** | **X** | **X** | **X** |  | **X** |  | **X** | **X** | **X** |
| **John Frater** | 38* | **X** | **X** | **X** | **X** | **X** | **X** | **X** | **X** |  | **X** | **X** | **X** | **X** | **X** |

### Breakdown of numbers of participant samples used in immunology assays

For the immunological analyses:

**Baseline (33 participants)**

33 of 34 recruited participants were included at baseline, prior to bNAb dosing and ATI (one participant was removed due to persistently high ‘background’ levels in the assays).

**Week 12 (26 participants)**

Of the 33 at baseline, seven participants were subsequently excluded from the Week 12 immunological analysis (four rebounded; two withdrew before Week 12; one high background).

**Week 20/24 (21 participants)**

Five of the 26 participants who reached Week 12 were excluded from the Week 20/24 analysis (four rebounded after Week 12 but before the week 20/24 visit, and one withdrew at week 15).

**Week 36 (17 participants)**

Of the 21 participants who were undetectable at Week 20/24, 4 experienced viral rebound prior to reaching Week 36 and were excluded, leaving 17 at Week 36.

**Second Dose**

Of 34 participants, 17 (50%) received a second dose of dual LS-bNAbs.

**Single Cell Analyses**

Participant samples used for single cell analysis: P8, P27, P15, P24, P21, P32

### Supplementary Table 1. bNAbs sensitivity and HLA typing

| **P.ID** | **Doses of bNAbs** | **Prevalence of Baseline RAMS** | **HLA class I alleles** | | | | | |
| --- | --- | --- | --- | --- | --- | --- | --- | --- |
|  |  |  | **A1** | **A2** | **B1** | **B2** | **C1** | **C2** |
| P03 | 1 | Nil | 02:01 | 03:01 | 44:02 | 47:01 | 05:01 | 06:02 |
| P04 | 1 | Nil | 29:02 | 30:01 | 13:02 | 14:02 | 06:02 | 08:02 |
| P05 | 1 | >15%:10-1074 | 02:01 | 02:01 | 07:02 | 27:05 | 01:02 | 07:02 |
| P07 | 1 | Nil | 01:01 | 03:01 | 35:01 | 57:01 | 04:01 | 06:02 |
| P08 | 2 | NA | 03:01 | 30:01 | 08:01 | 13:02 | 06:02 | 07:02 |
| P15 | 2 | Nil | 02:01 | 68:01 | 35:02 | 51:08 | 04:01 | 15:02 |
| P17 | 2 | NA | 02:01 | 31:01 | 35:03 | 51:01 | 04:01 | 16:02 |
| P18 | 2 | NA | 24:02 | 26:01 | 27:05 | 35:01 | 02:02 | 04:01 |
| P19 | 1 | Nil | 02:01 | 25:01 | 39:01 | 57:01 | 06:02 | 12:03 |
| P21 | 2 | Nil | 11:01 | 24:02 | 40:01 | 57:01 | 03:04 | 07:01 |
| P22 | 1 | >15%:3BNC117 | 01:01 | 02:01 | 08:01 | 44:02 | 05:01 | 07:01 |
| P24 | 2 | Nil | 02:01 | 03:01 | 14:02 | 37:01 | 06:02 | 08:02 |
| P25 | 1 | Nil | 03:01 | 26:01 | 35:01 | 39:01 | 04:01 | 12:03 |
| P27 | 2 | Nil | 02:01 | 03:01 | 15:16 | 18:01 | 05:01 | 14:02 |
| P31 | 1 | Nil | 02:01 | 24:02 | 35:01 | 40:01 | 03:04 | 08:01 |
| P32 | 2 | Nil | 02:01 | 02:01 | 07:02 | 44:03 | 07:02 | 16:01 |
| P33 | 1 | Nil | 02:01 | 11:01 | 15:03 | 51:01 | 12:03 | 15:02 |
| P37 | 1 | Nil | 03:01 | 11:01 | 15:01 | 35:01 | 03:04 | 04:01 |
| P39 | 2 | Nil | 01:01 | 68:01 | 08:01 | 51:01 | 07:01 | 15:02 |
| P40 | 2 | <15%:10-1074 | 01:01 | 02:01 | 07:02 | 44:02 | 05:01 | 07:02 |
| P41 | 1 | >15%:3BNC117 | 02:01 | 11:01 | 30:01 | 51:01 | 12:03 | 15:02 |
| P44 | 2 | >15%:3BNC117 | 01:01 | 02:01 | 35:01 | 57:01 | 04:01 | 06:02 |
| P46 | 2 | Nil | 02:01 | 24:02 | 15:18 | 52:01 | 07:02 | 07:04 |
| P48 | 2 | Nil | 03:01 | 11:01 | 40:01 | 55:01 | 03:03 | 03:04 |
| P50 | 2 | NA | 03:01 | 32:01 | 44:02 | 44:02 | 05:01 | 07:04 |
| P51 | 1 | Nil | 02:01 | 25:01 | 07:02 | 15:01 | 03:03 | 07:02 |
| P54 | 1 | NA | 03:01 | 68:01 | 07:02 | 45:01 | 06:02 | 07:02 |
| P55 | 1 | >15%:3BNC117 | 02:01 | 02:01 | 07:10 | 44:02 | 05:01 | 07:02 |
| P57 | 2 | Nil | 29:02 | 33:01 | 14:02 | 52:01 | 08:02 | 12:02 |
| P59 | 2 | Nil | 02:01 | 03:01 | 27:05 | 51:01 | 01:02 | 15:02 |
| P62 | 2 | <15%:3BNC117 | 02:01 | 02:01 | 15:01 | 44:05 | 02:02 | 03:28 |
| P63* | 1 | Nil | 01:01 | 03:01 | 07:02 | 08:01 | 07:01 | 07:01 |
| P65 | 1 | Nil | 02:01 | 02:01 | 44:02 | 44:02 | 05:01 | 05:01 |
| P67 | 1 | <15%:10-1074  <15%:3BNC117 | 03:01 | 23:01 | 44:02 | 51:01 | 07:04 | 16:01 |

Abbreviations: RAMS, Resistance Associated Mutations; bNAb, broadly neutralizing antibodies; HLA, human leukocyte antigen; NA, not available; * High background and not used.

### Supplementary Table 2. Flow Cytometry Panels

Phenotyping Assay Antibody Panels

| Cell surface antibodies |  |  |
| --- | --- | --- |
| anti-CXCR3 | APC | (clone 1C6, BD Biosciences, San Jose, USA) |
| anti-CCR7 | PE | (clone G043H7, BioLegend, San Diego, USA), |
| anti-CD3 | BV711 | (clone UCHT1, BioLegend, San Diego, USA), |
| anti-CD127 | BV605 | (clone A019D5, BioLegend, San Diego, USA), |
| anti-HLA-DR | APC-Cy7 | (clone L243, BioLegend, San Diego, USA), |
| anti-CD38-PE | Texas Red | (clone HIT2, BD Biosciences, San Jose, USA) |
| anti-CD4 | BV570 | (clone RPAT4, BioLegend, San Diego, USA), |
| anti-CXCR5 | BB515 | (clone RF8B2, BD Biosciences, San Jose, USA), |
| anti-PD1 | BV421 | (clone EH12.2H7, BioLegend, San Diego, USA), |
| anti-CD45RA | BV650 | (clone HI100, BioLegend, San Diego, USA), |
| anti-CCR6 | BB700 | (clone 11A9, BD Biosciences, San Jose, USA), |
| anti-CD8 | APC-R700 | (clone RPA-T8, BD Biosciences, San Jose, USA), |
| anti-CD25 | PE-Cy7 | (clone 2A3, BD Biosciences, San Jose, USA), |
| anti-CD45RA | BV650 | (clone HI100, BioLegend, San Diego, USA), |
| live-dead aqua |  | (Invitrogen, MA, USA), |
| anti-CD19 (dump) | BV510 | (clone HIB19, BioLegend, San Diego, USA), |
| anti-CD14 (dump) | BV510 | (clone M5E2, BioLegend, San Diego, USA), a |
| anti-TIGIT | PerCP-EF710 | (clone GIGD7, Invitrogen, MA, USA), |
| anti-CCR7 | BV650 | (clone HI100, BioLegend, San Diego, USA), |
| anti-LAG-3 | PE-Cy7 | (clone 11C3C65, BioLegend, San Diego, USA), |
| anti-TIM3 | PE | (clone F38-2E2, BioLegend, San Diego, USA), |
| anti-CD45RA | BV711 | (clone HI100, BioLegend, San Diego, USA), |
| anti-CXCR5 | AF647 | (clone RF8B2, BD Biosciences, San Jose, USA), |
| anti-CD3 | BV605 | (clone UCHT1, BioLegend, San Diego, USA), |
| Intracellular Antibodies |  |  |
| anti-Tbet | FITC | (clone 4B10, BioLegend, San Diego, USA) |
| anti-Eomes | PE-Texas Red | (clone WD1928, Life Technologies, CA, USA) |

AIM Assay Antibody Panels

| Pre-Stim |  |  |
| --- | --- | --- |
| anti-CXCR3 | APC | (clone 1C6, BD Biosciences, San Jose, USA) |
| Post-Stim and Fc Block |  |  |
| anti-CD3 | BV605 | (clone UCHT1, BioLegend, San Diego, USA) |
| anti-CD4 | BV570 | (clone RPA-T4, BioLegend, San Diego, USA) |
| anti-CD8 | APC-R700 | (clone RPA-T8, BD Biosciences, San Jose, USA) |
| anti-CD134 (OX40) | PE | (clone L106, BD Biosciences, San Jose, USA) |
| anti-CXCR5 | BB515 | (clone RF8B2, BD Biosciences, San Jose, USA) |
| anti-CCR6 | APC-Cy7 | (clone G034E3, BioLegend, San Diego, USA) |
| anti-PD-1 | BV421 | (clone EH12.2H7, BioLegend, San Diego, USA) |
| anti-CD137 | PECF-594 | (clone 4B4-1, BioLegend, San Diego, USA) |
| anti-CD39 | PerCP-Efluor710 | (clone eBioA1, Invitrogen, MA, USA) |
| anti-CD25 | PE-Cy7 | (clone 2A3, BD Biosciences, San Jose, USA) |
| anti-CD69 | BV650 | (clone FN50, BioLegend, San Diego, USA), |
| live-dead aqua |  | (Invitrogen, MA, USA) |
| anti-CD19 (dump) | BV510 | (clone HIB19, BioLegend, San Diego, USA) |
| anti-CD14 (dump) | BV510 | (clone M5E2, BioLegend, San Diego, USA) |

Proliferation Assay Antibody Panels

| anti-CD3 | FITC | (clone HIT3a, BD Biosciences, San Jose, USA) |
| --- | --- | --- |
| anti-CD4 | APC | (clone RPA-T4, BD Biosciences, San Jose, USA) |
| anti-CD8 | PE-Cy7 | (clone SK1, BioLegend, San Diego, USA) |

Supplementary Figures

### Supplementary Figure 1. Gamma interferon ELISpot format – matrix

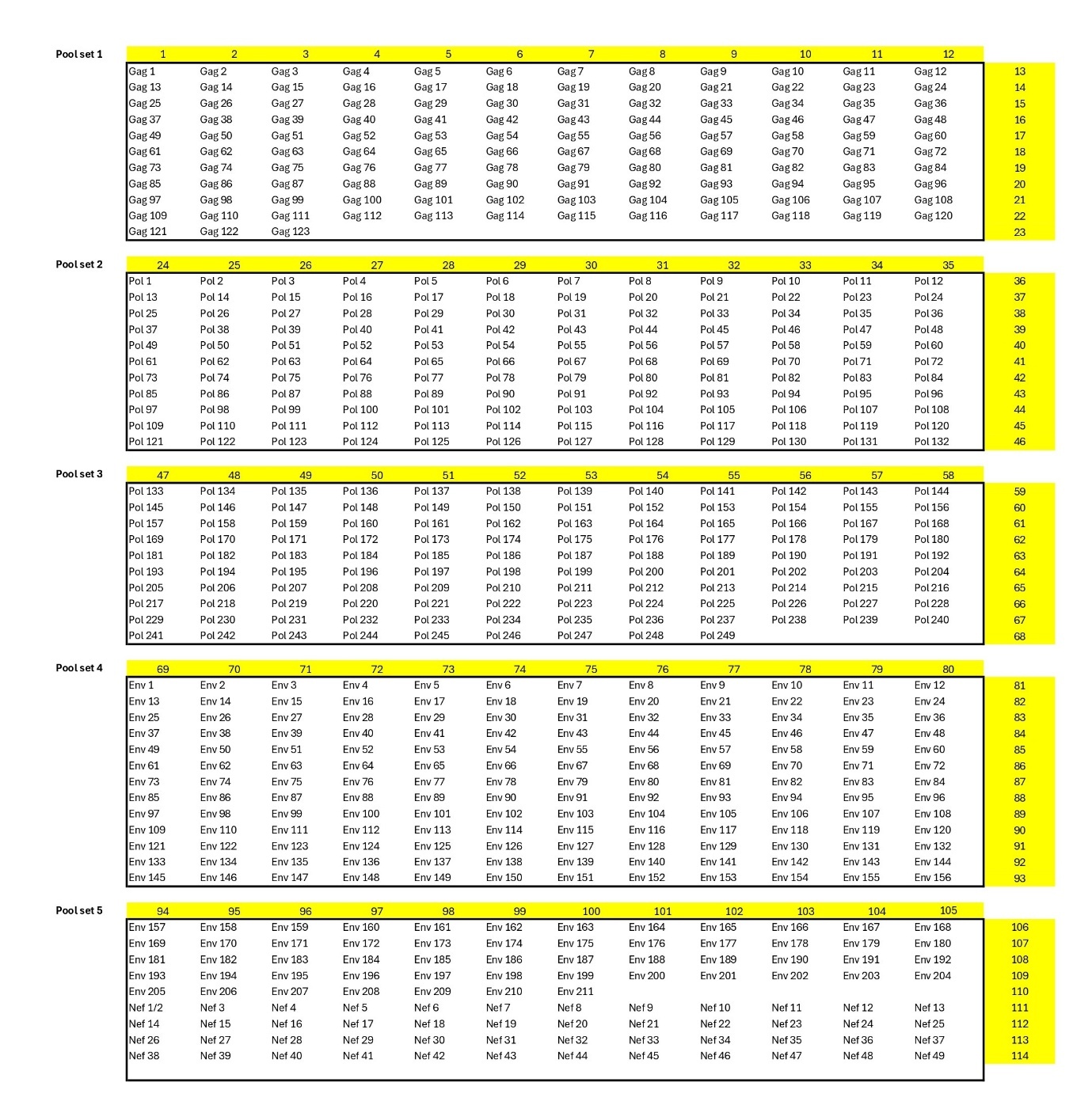

Breakdown of the 114 pools use to screen HIV-specific responses across the HIV Gag, Pol, Env and Nef proteins. The matrix layout ensures every peptide is presented once across two pools, allowing deconvolution of individual responses as well as total protein-specific responses. Peptide numbering is based on the NIH HIV Reagent Program: https://www.niaid.nih.gov/research/nih-hiv-reagent-program

### Supplementary Figure 2. Immune responses to Gag, Pol, Env and Nef

a. Gag Responses for Participants remaining undetectable for 36 weeks

b. ELISpot data

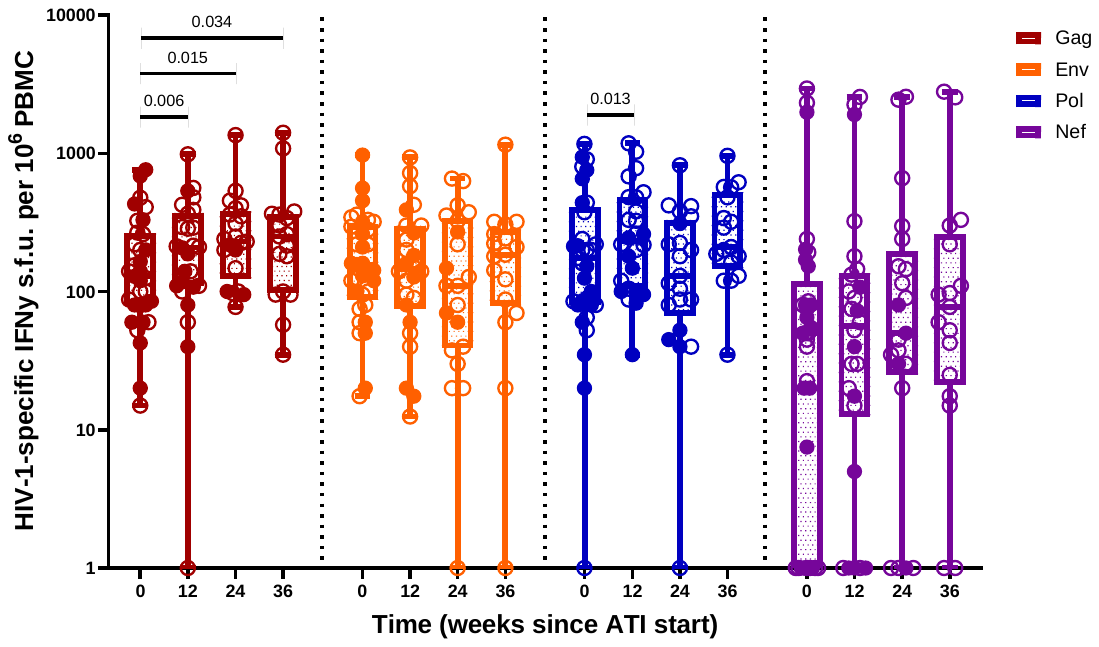

c. AIM data

**CD4+ T cell responses to HIV Gag, Env, Pol and Nef for 36 weeks after bNAbs and ATI**

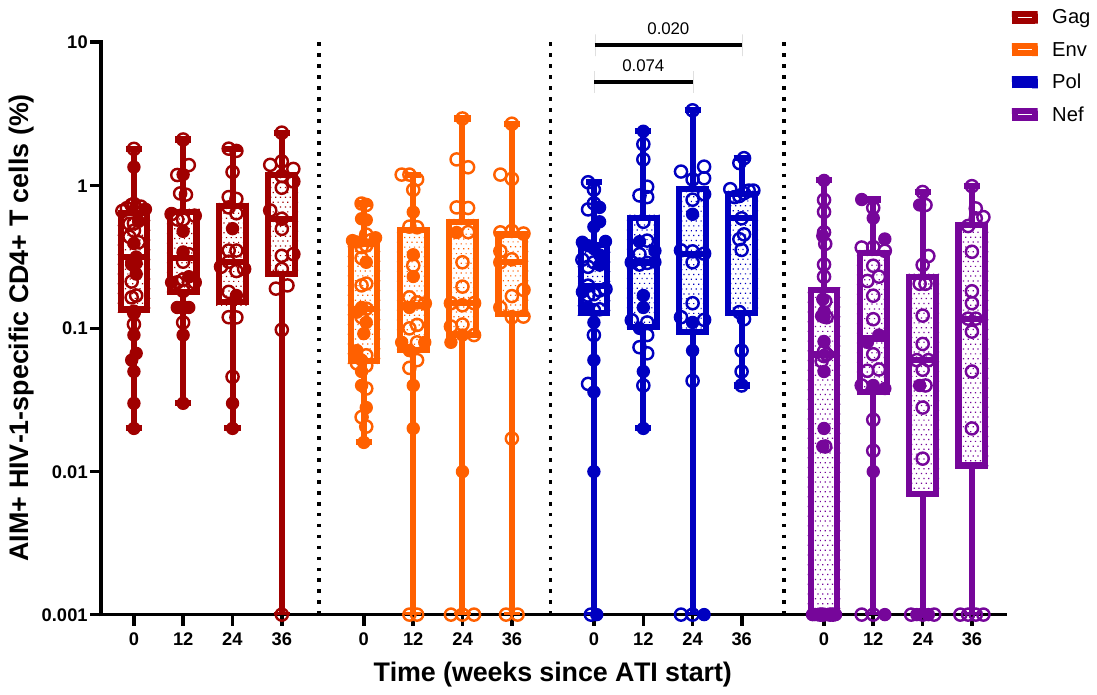

**CD8+ T cell responses to HIV Gag, Env, Pol and Nef for 36 weeks after bNAbs and ATI**

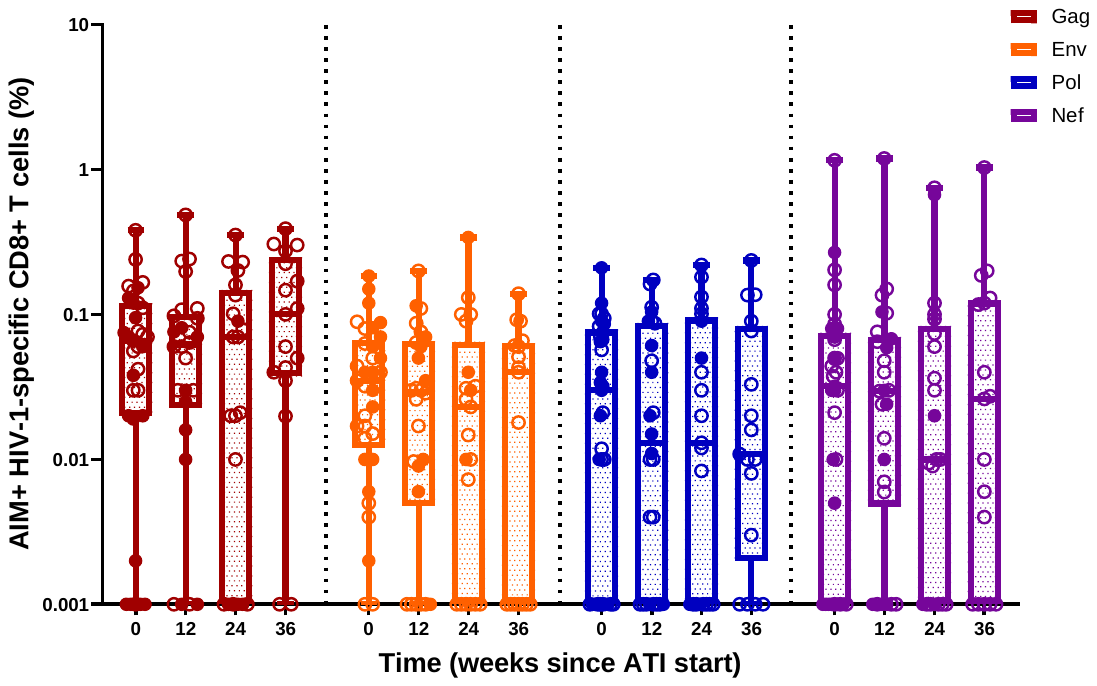

**d.** Proliferation data

**CD4+ T cell responses to HIV Gag, Env, Pol and Nef for 36 weeks after bNAbs and ATI**

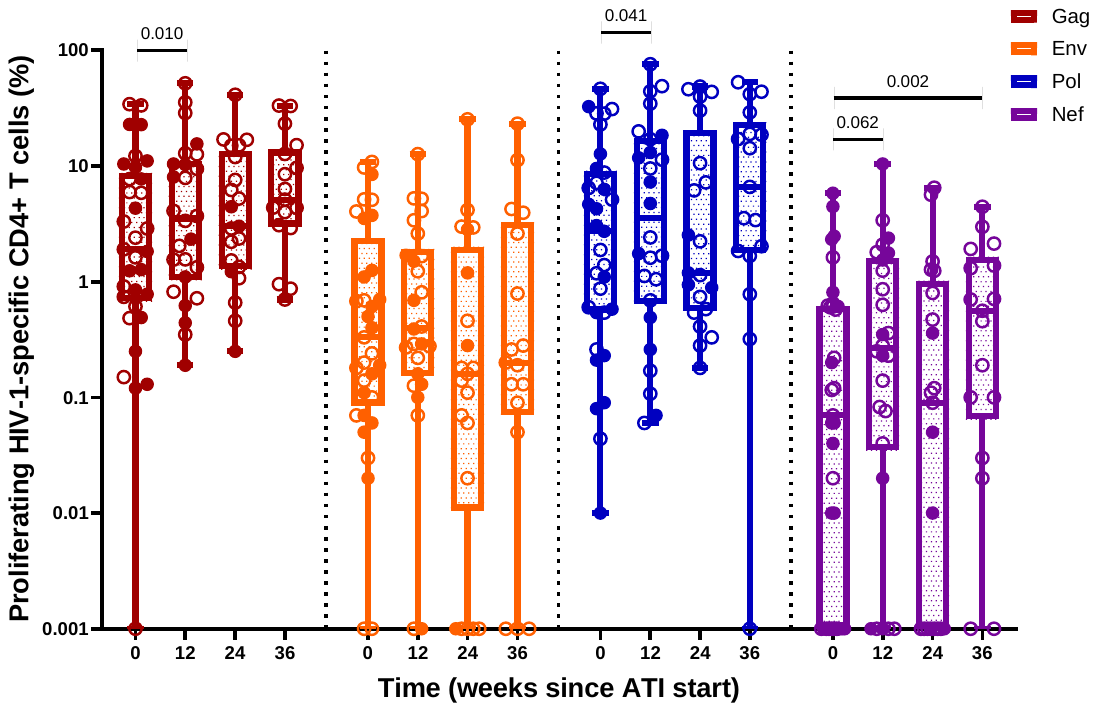

**CD8+ T cell responses to HIV Gag, Env, Pol and Nef for 36 weeks after bNAbs and ATI**

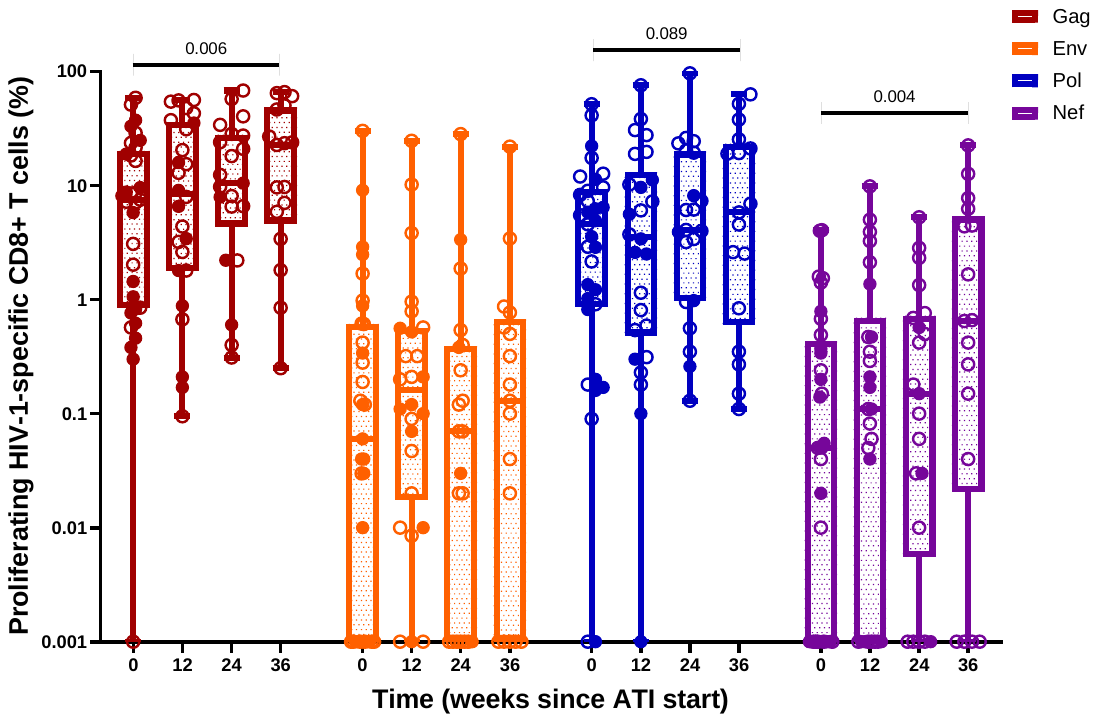

### Supplementary Figure 3. T cell phenotyping – full dataset

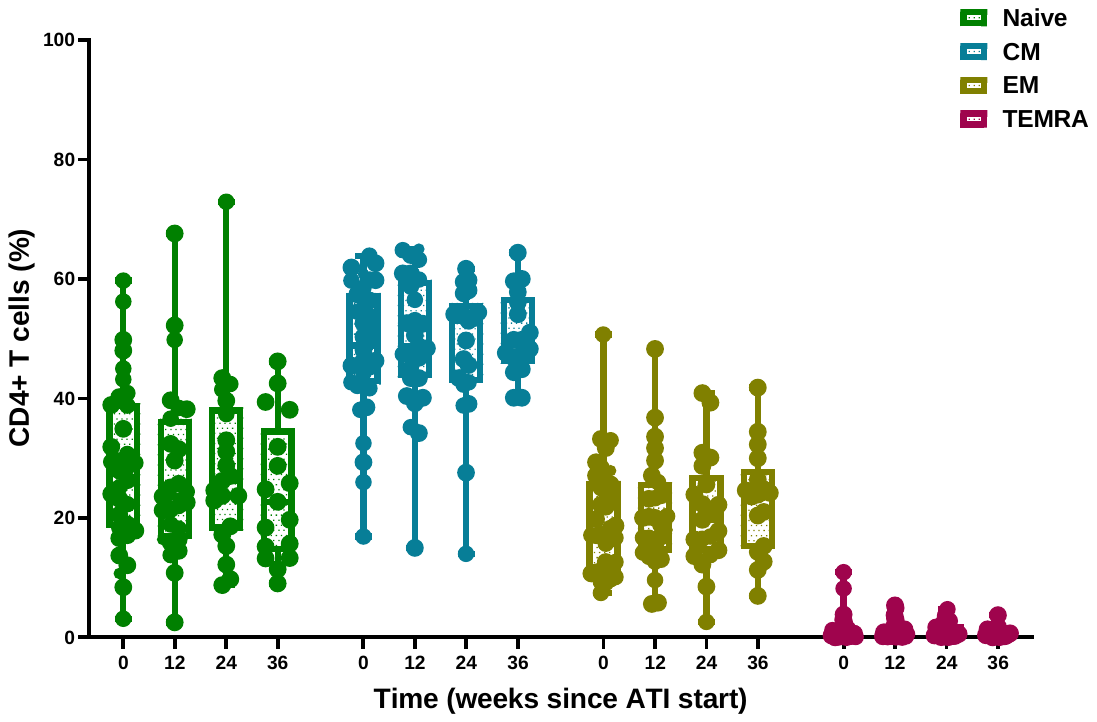

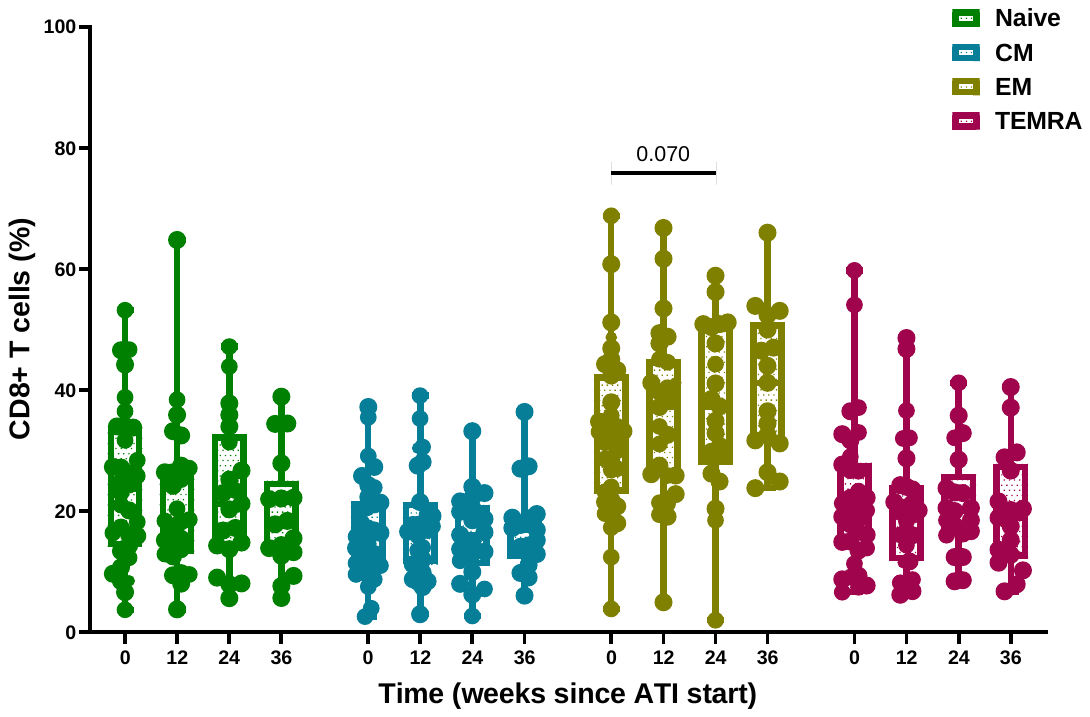

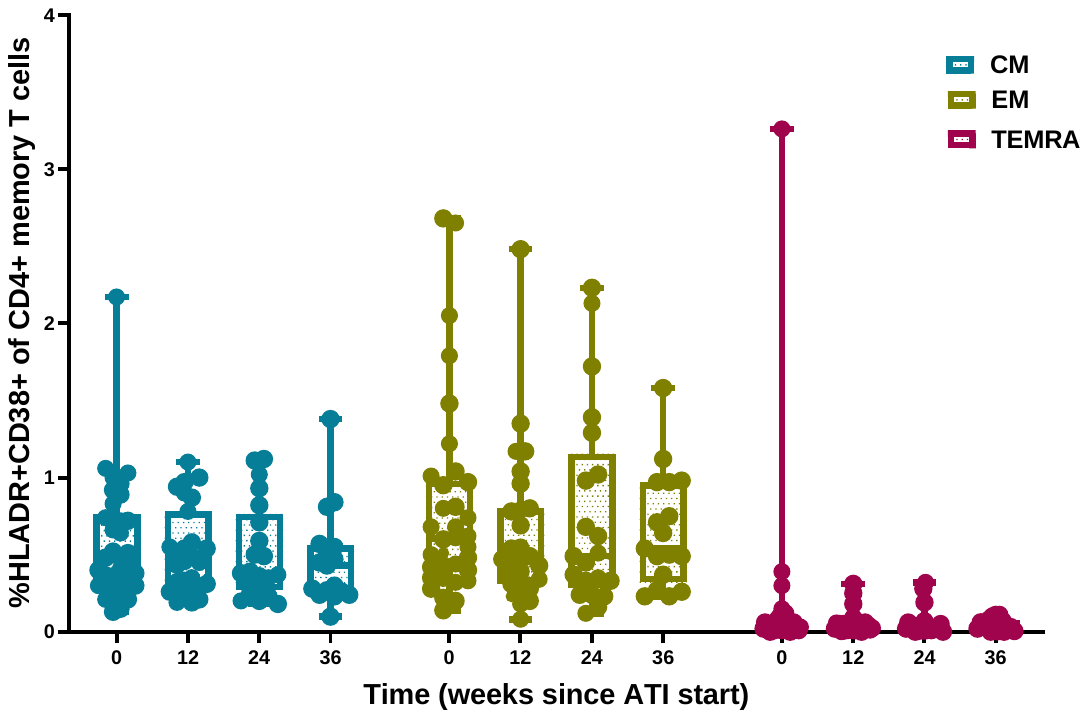

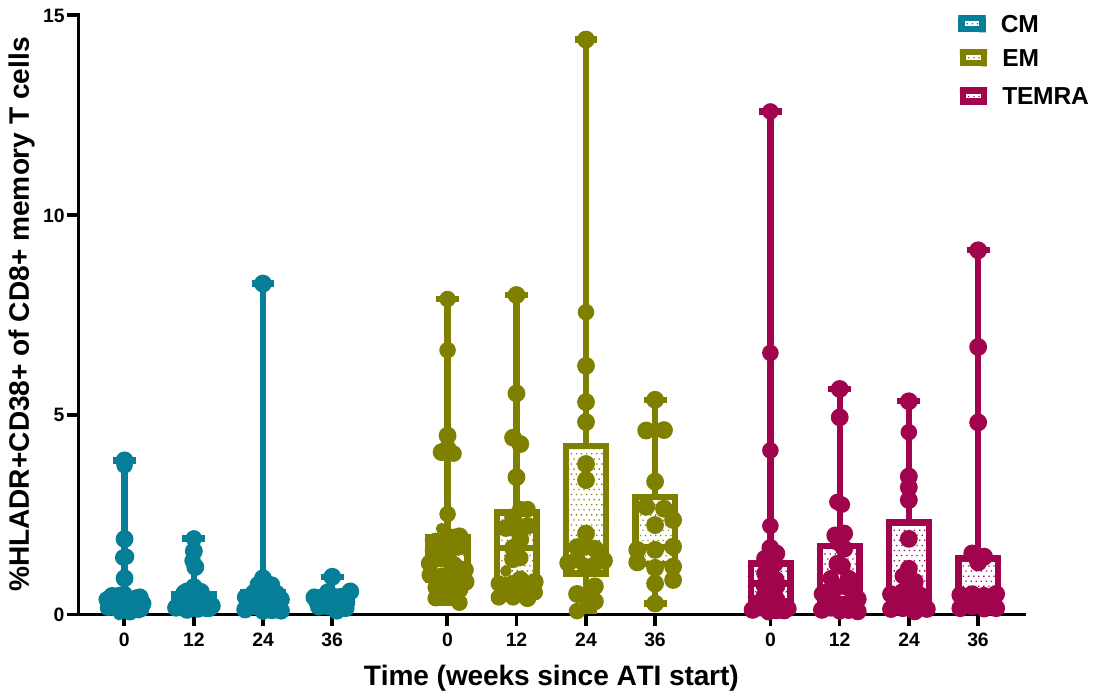

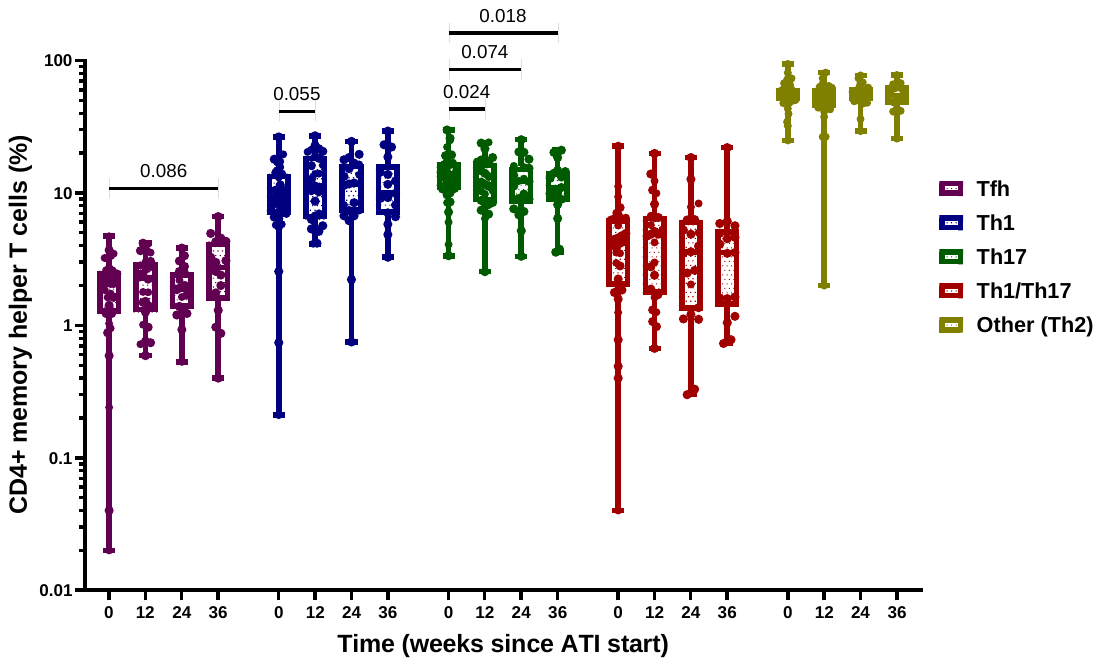

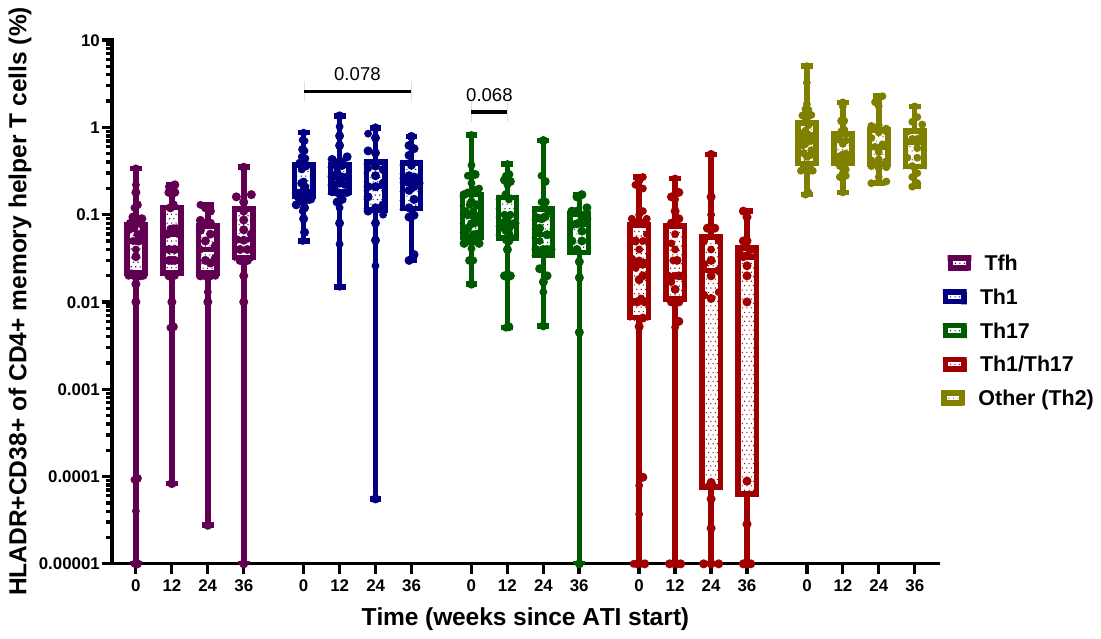

### Supplementary Figure 4. Exhaustion markers – full dataset

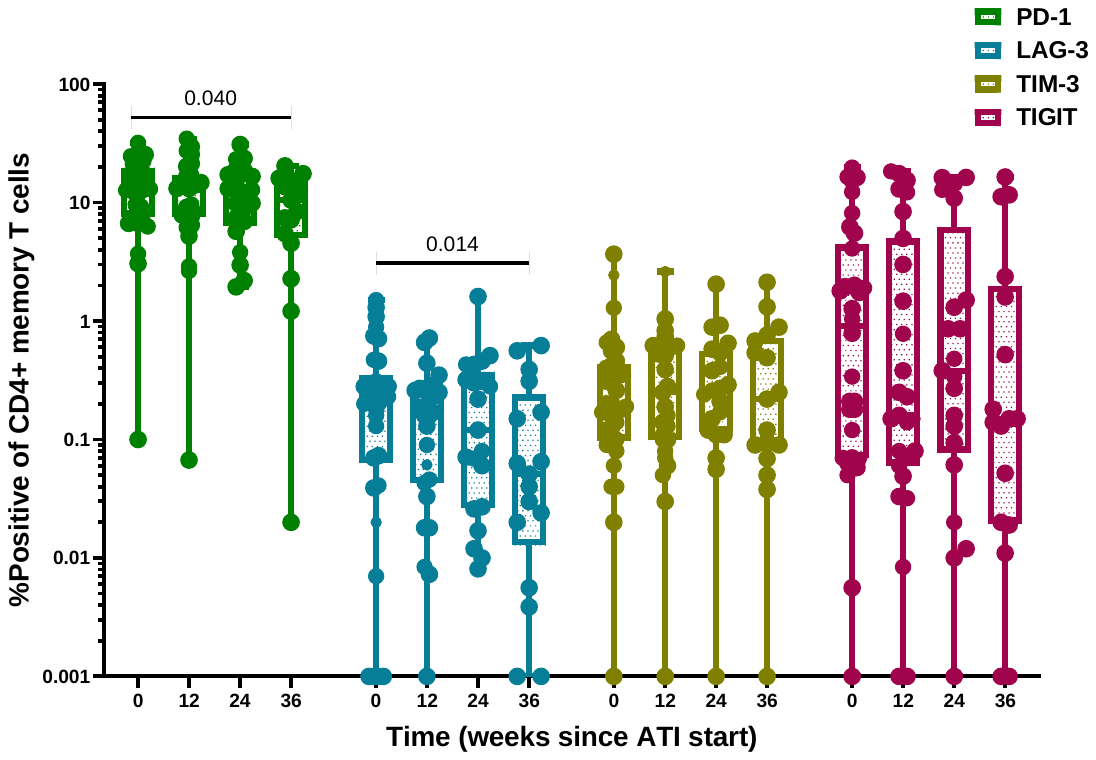

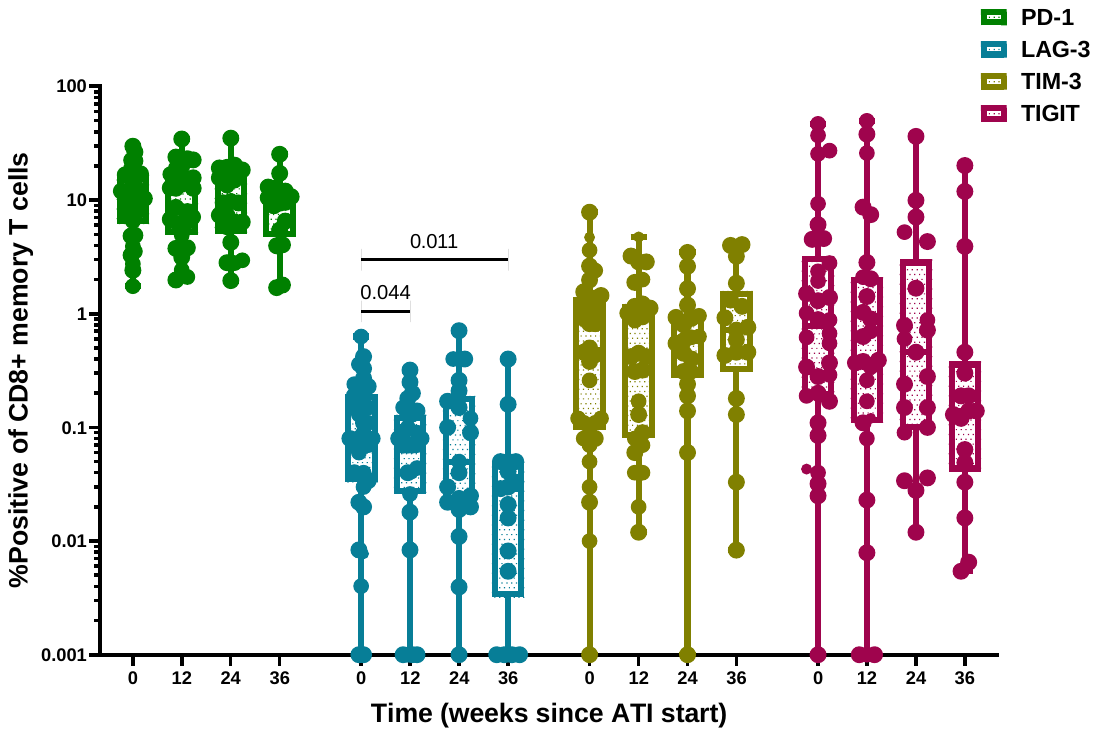

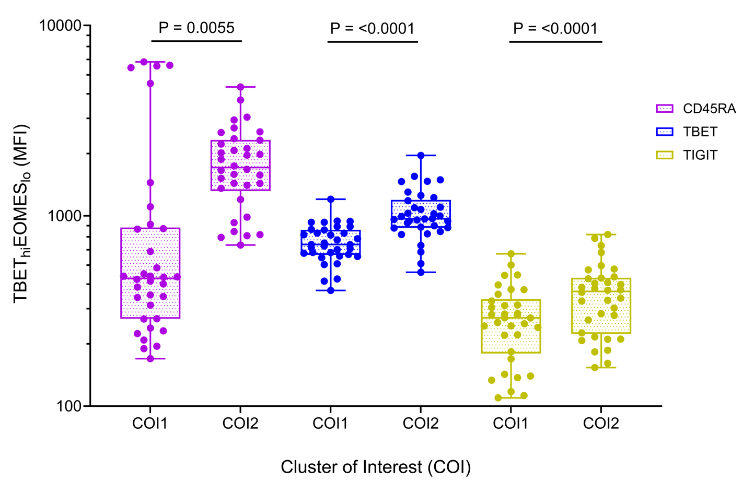

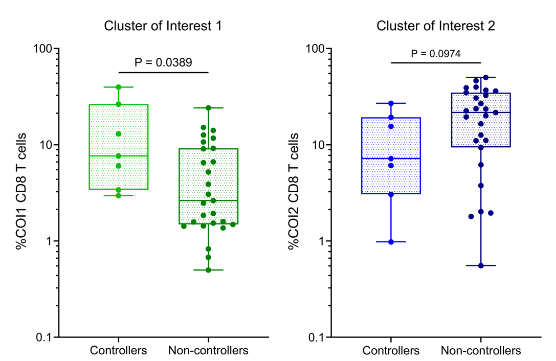

### Supplementary Figure 5. Cox models for baseline predictors of time to rebound

**Baseline responses for participants who contained no resistance associated mutations (RAMs) in Env using rebound ‘time from last dose’**

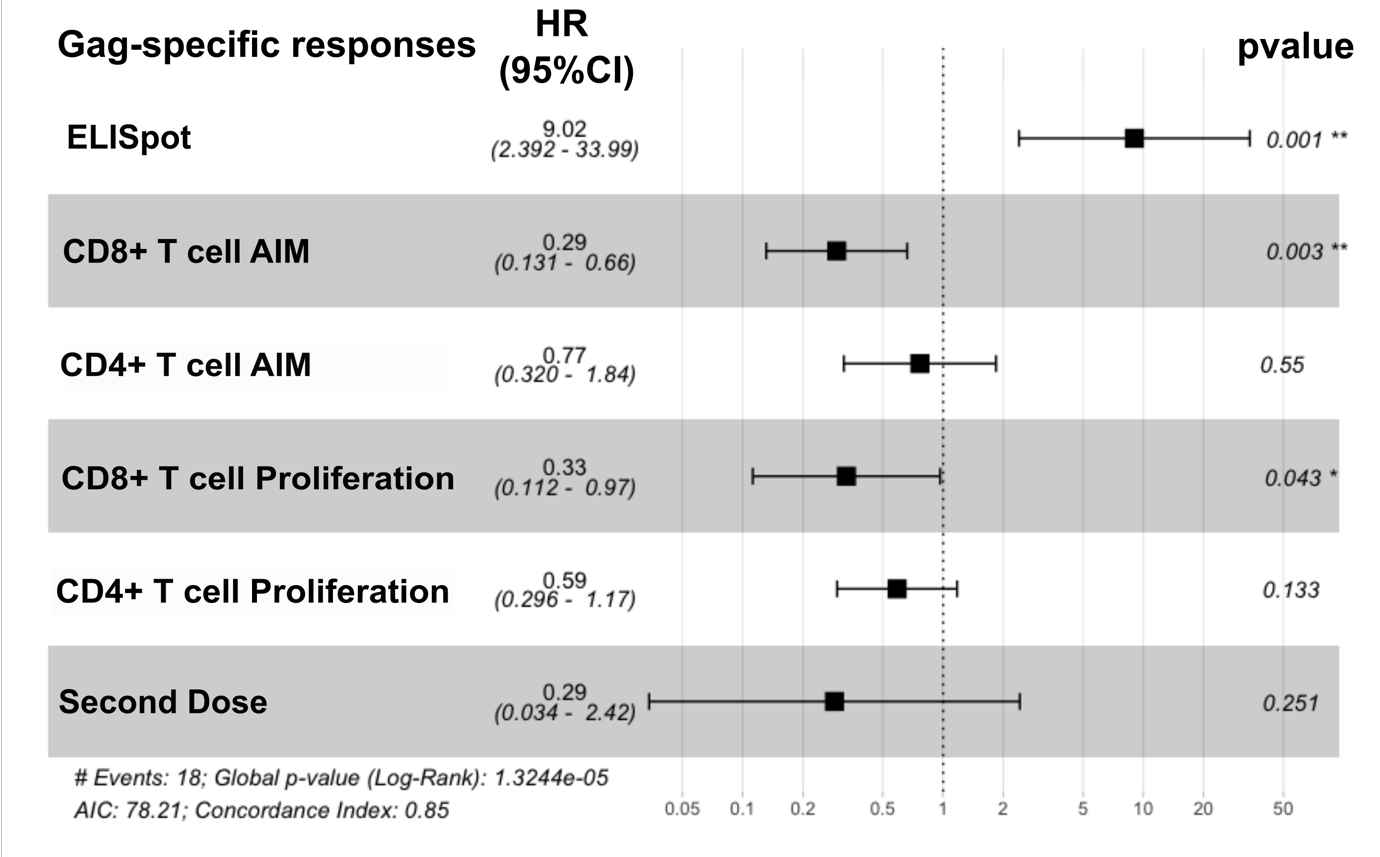

### Supplementary Figure 6. Cox models for fold-change predictors of time to rebound

a. Fold change: Weeks 12-24; All participants

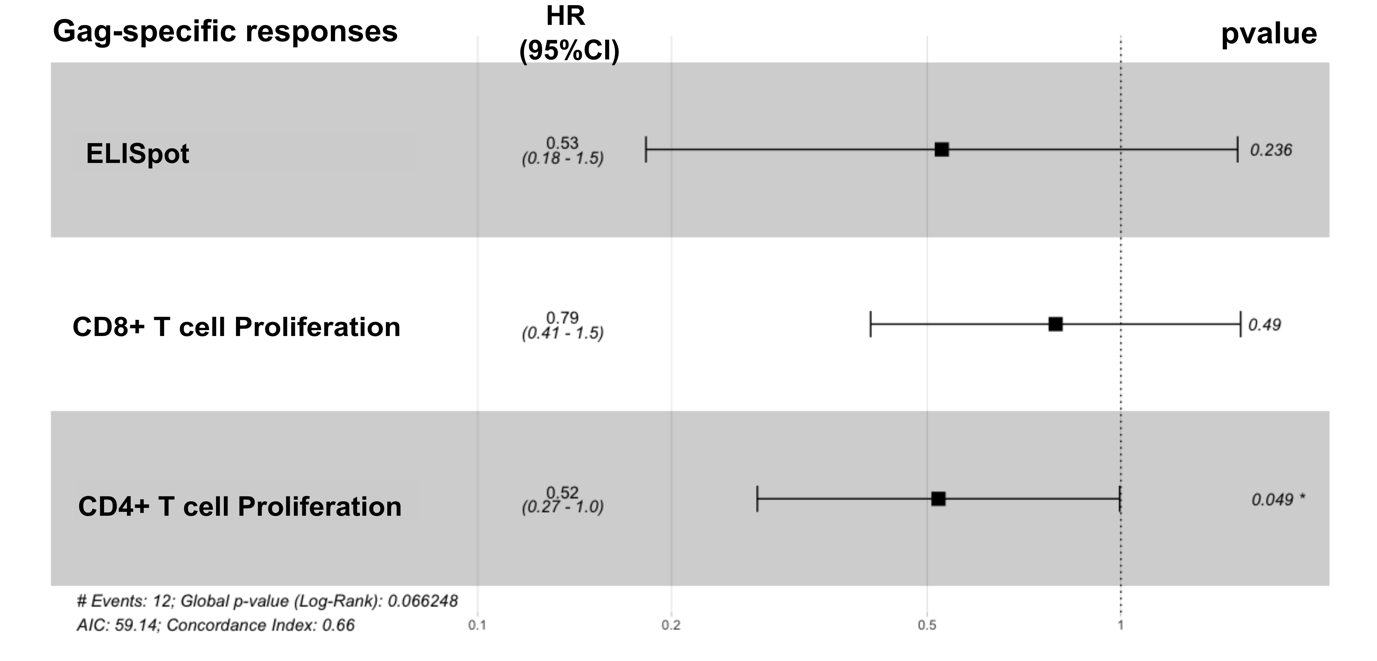

b. Fold change: Weeks 0 to 24; All participants

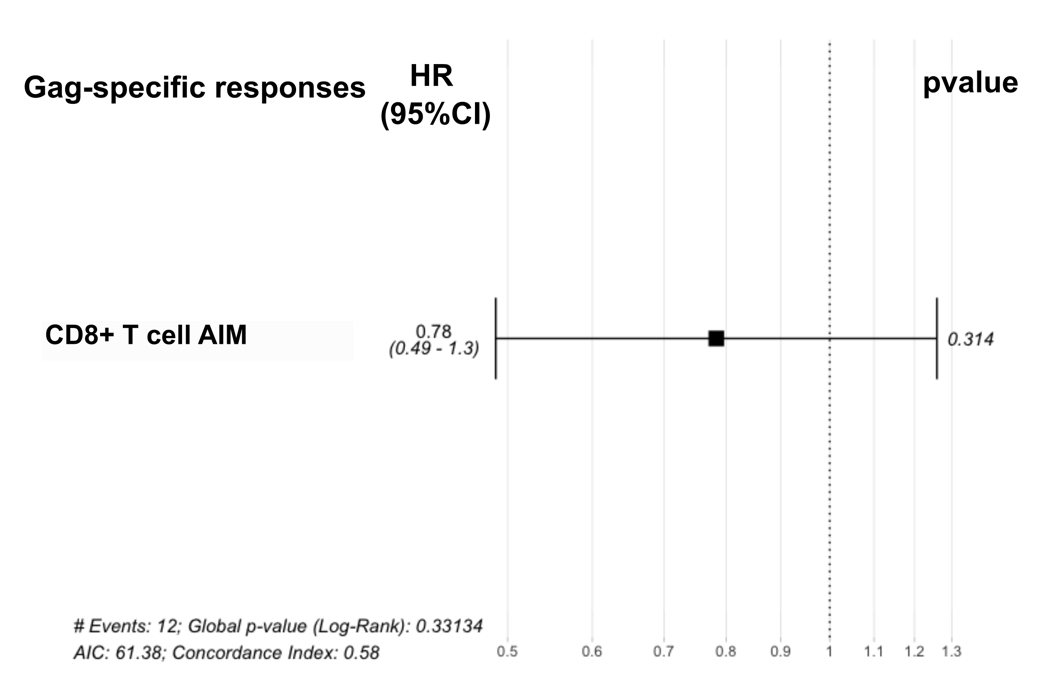

### Supplementary Figure 7. Time-dependent Cox models of HLA associations with virological outcomes

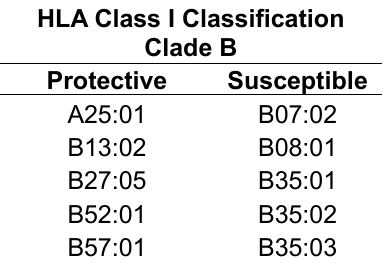

a.

b. **Hazard Score of all participants**

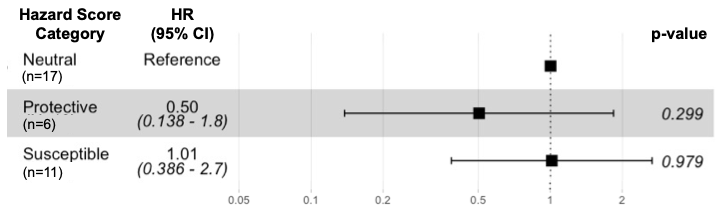

c. **Participants without baseline RAMs**

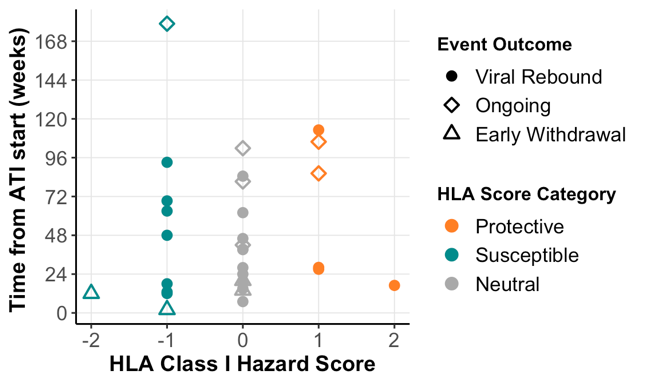

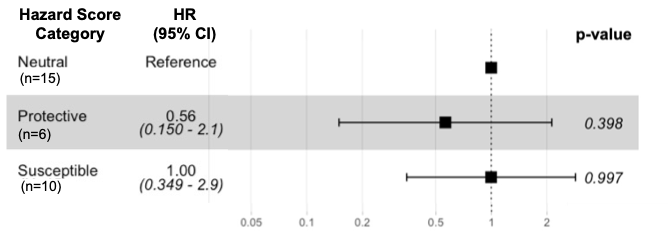

d. **Time to rebound using discrete categorical variables**

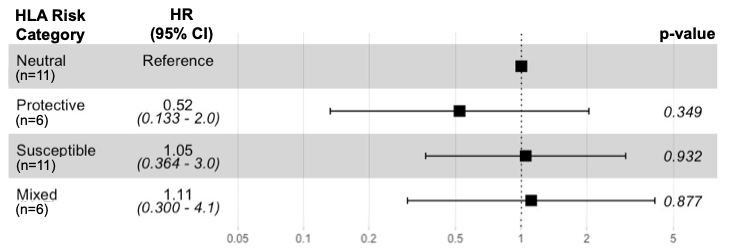

### Supplementary Figure 8. CMV responses across different assays and timepoints

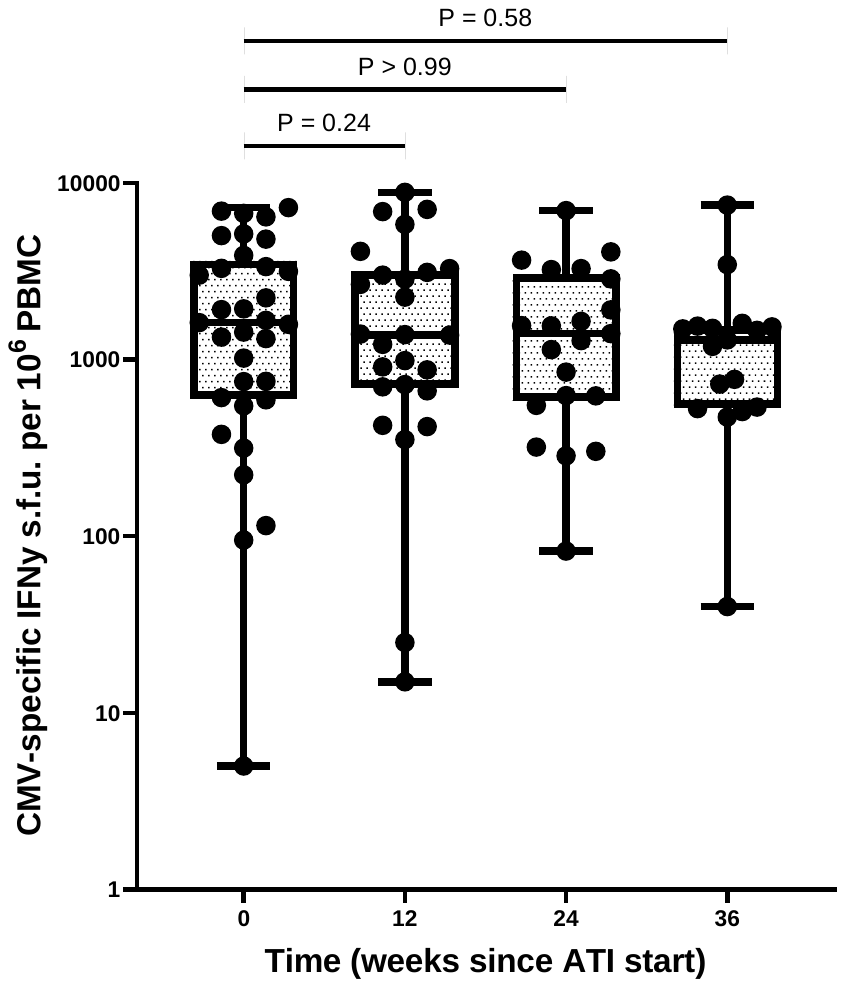

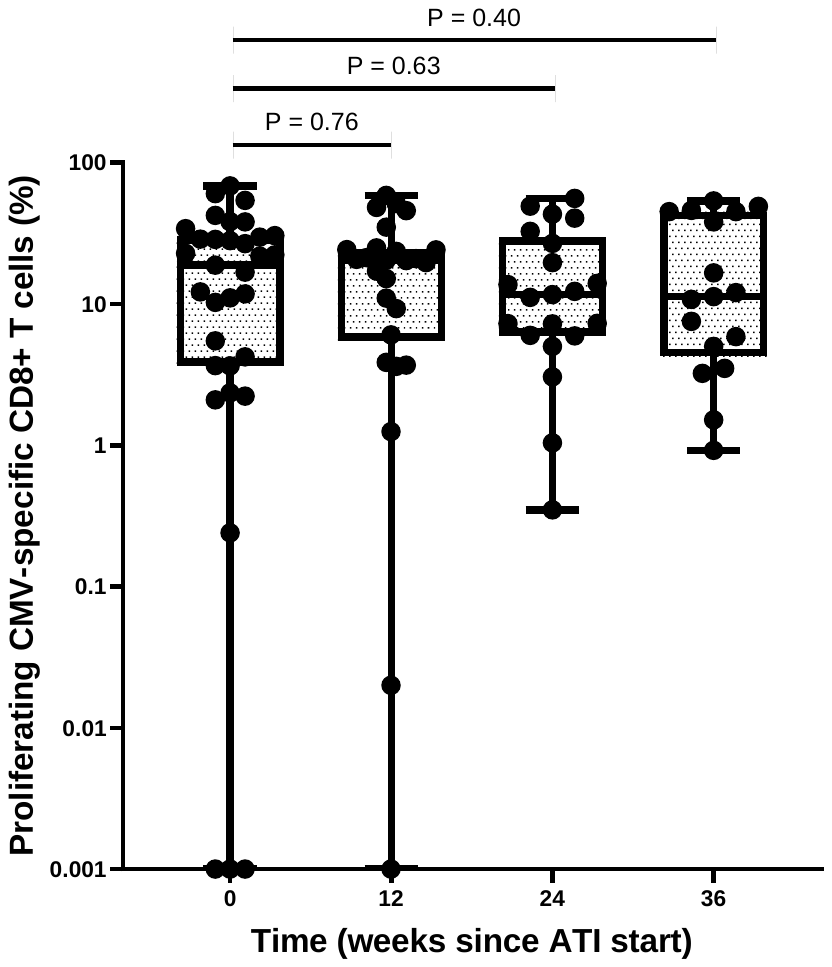

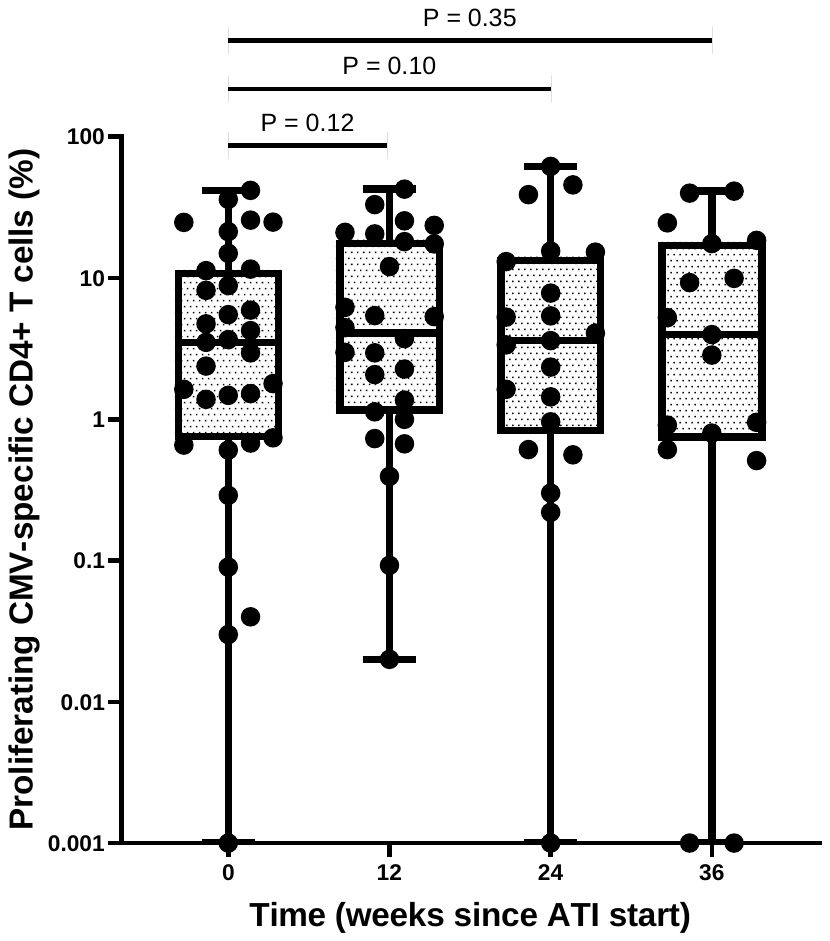

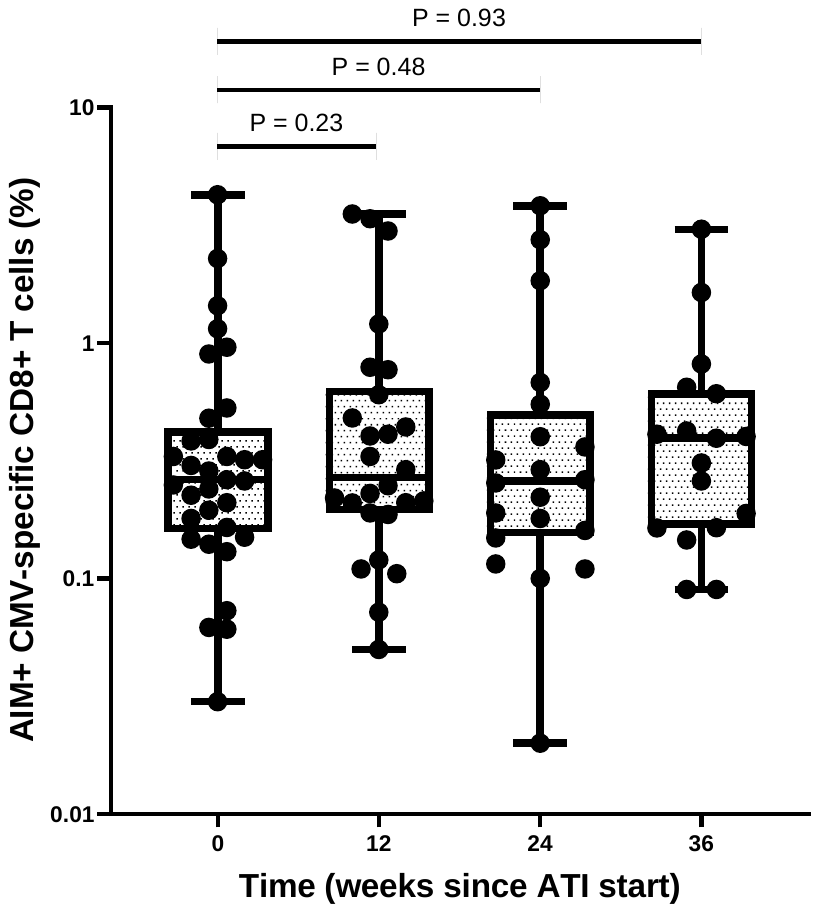

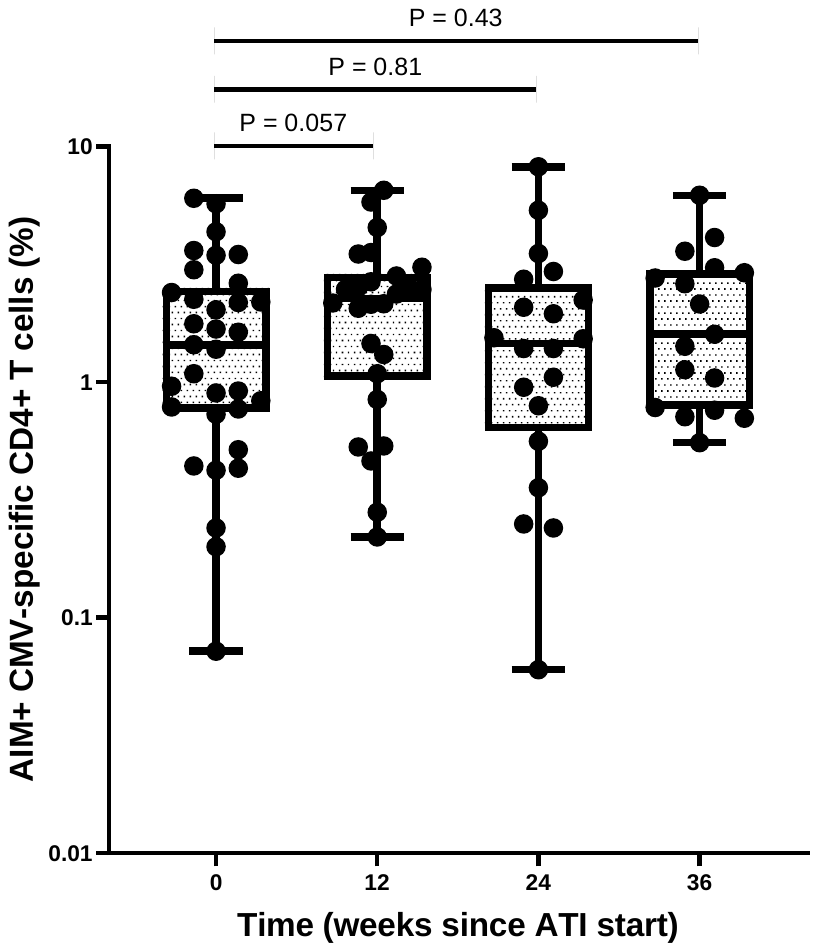
